# Polygenic resilience scores capture protective genetic effects for Alzheimer’s disease

**DOI:** 10.1101/2022.05.10.22274858

**Authors:** Jiahui Hou, Jonathan L. Hess, Nicola Armstrong, Joshua C. Bis, Benjamin Grenier-Boley, Ida K. Karlsson, Ganna Leonenko, Katya Numbers, Eleanor K. O’Brien, Alexey Shadrin, Anbupalam Thalamuthu, Qiong Yang, Ole A. Andreassen, Henry Brodaty, Margaret Gatz, Nicole A. Kochan, Jean-Charles Lambert, Simon M. Laws, Colin L. Masters, Karen A. Mather, Nancy L. Pedersen, Danielle Posthuma, Perminder S. Sachdev, Julie Williams, the Alzheimer’s Disease Neuroimaging Initiative, Chun Chieh Fan, Stephen V. Faraone, Christine Fennema-Notestine, Shu-Ju Lin, Valentina Escott-Price, Peter Holmans, Sudha Seshadri, Ming T. Tsuang, William S. Kremen, Stephen J. Glatt

## Abstract

Polygenic risk scores (PRSs) can boost risk-prediction in late-onset Alzheimer’s disease (LOAD) beyond apolipoprotein E (*APOE)* but have not been leveraged to identify genetic resilience factors. Here, we sought to identify resilience-conferring common genetic variants in 1) unaffected individuals having high PRSs for LOAD, and 2) unaffected *APOE*-_ε_4 carriers also having high PRSs for LOAD. We used genome-wide association study (GWAS) to contrast “resilient” unaffected individuals at the highest genetic risk for LOAD with LOAD cases at comparable risk. From GWAS results, we constructed polygenic resilience scores to aggregate the addictive contributions of risk-orthogonal common variants that promote resilience to LOAD. Replication of resilience scores was undertaken in eight independent studies. We successfully replicated two polygenic resilience scores that reduce genetic-risk penetrance for LOAD. We also showed that polygenic resilience scores positively correlate with polygenic risk scores in unaffected individuals, perhaps aiding in staving off disease. Our findings align with the hypothesis that a combination of risk-independent common variants mediates resilience to LOAD by moderating genetic disease risk.

## 1. Introduction

Alzheimer’s disease (AD) is the leading cause of dementia^1^. AD exists as two genetically distinct forms: early-onset AD, which is caused by autosomal dominant mutations in genes (*PSEN1*; *PSEN2*; *APP; SORL1*) and typically has an onset of symptoms between the ages of 40 to 60 years^2^, and the more common late-onset AD (LOAD), which is sporadic, polygenic, and typically has an onset of symptoms in the mid-60s^3^. Elevated risk of LOAD is associated with a host of lifestyle factors and medical conditions, such as a high-fat diet, heavy drinking and smoking, cardiovascular disease, type-2 diabetes, and traumatic brain injury^4^. More importantly, the heritability of LOAD from twin studies was estimated at 58%-79%^5^, and its estimates from single nucleotide polymorphisms (SNPs) range from 13% to 33%^6-9^. The goal of this study is to determine whether genes also play a role in resilience to LOAD. We used an innovative approach first introduced and applied in schizophrenia as a general framework for resilience research^10^, focusing on individuals at the highest levels of genetic risk.

To date, genome-wide association studies (GWASs) have discovered close to 50 genome-wide significant loci (*p*<5*e*-08) associated with LOAD risk^9, 11-20^. The _ε_4 allele of apolipoprotein E (*APOE*) is the polymorphism with the strongest effect on LOAD susceptibility^21^. Beyond *APOE-*_ε_4, there may be thousands of additional genetic polymorphisms that make small individual contributions to the overall risk for LOAD^22-25^. A polygenic risk score (PRS)^26^ can be derived by summing the weighted effect of SNPs to identify a single genetic risk variable that reflects one’s relative susceptibility to LOAD. Recent LOAD PRSs capture most of the SNP-heritability for LOAD^9, 24, 27^. Extensive research shows that PRSs boost the accuracy of LOAD diagnosis beyond the performance of *APOE*^22-25^, and capture LOAD phenotypic variability not explained by *APOE* status^28, 29^.

Revealing the genetic architecture of LOAD is vital for understanding its etiology and identifying molecular targets for innovative therapeutic interventions. Yet, knowledge of risk factors might be fruitfully complemented by an understanding of resilience-associated or - promoting mechanisms as well. As such, some AD research has shifted focus from symptomatic cases to healthy aging individuals or asymptomatic individuals at elevated risk^30^. This was motivated by the premise that high-risk asymptomatic individuals, yet unaffected, may provide clues that protect them against AD. Here, we employ the term “resilience” to indicate individuals who show better than expected outcomes in the face of high genetic risk for disease^30-35^.

Increasing evidence suggests that several factors—including education, literacy, physical activity, and mental activity—can moderate the risk for LOAD^31, 32, 36^, and it is estimated that one-third to 40% of dementia cases might be preventable^36, 37^. These moderation effects may be explained by reverse causation^38^, but genetic influences—which are not subject to reverse causation—also underlie these factors. Educational attainment^39, 40^ and, particularly, general cognitive ability^40, 41^ are heritable. Thus, some of these factors may also confer resilience-enhancing genetic effects. Notably, some genetic variants, such as *APOE*-_ε_2^42^ and the *APP* A673T variant^43^, have been identified as protective for LOAD. However, the biological mechanisms that drive the protective effects remain largely unknown. Importantly, we consider such protective effects to be fundamentally different from the “resilience” effects we sought in our study, in that protective factors are generally operative across the full range of risk, whereas resilience factors are only operative in those at the highest risk for disease. Very little work has been aimed at identifying additional genetic resilience factors that potentially moderate the genetic risk established by the cumulative effects of risk-associated alleles and their corresponding protective alleles. Genetic resilience against risk for LOAD has been investigated through diverse approaches based on varying conceptualizations and measurements used to identify individuals at high risk. As aggregation of beta-amyloid plaques and tau tangles in the brain are two of the neuropathological hallmarks of LOAD^44^, a principal focus of resilience has been on asymptomatic individuals who have cognition levels that are better than predicted based on these pathologies^45-47^. Other studies have leveraged known genetic risk factors to study resilience. For example, in *APOE*-_ε_4 carriers, over a dozen SNPs have been reported to potentially facilitate resilience, such as rs10553596 in *CASP7*^48^ and the rs4934 nonsynonymous variant in *SERPINA3*^49, 50^. However, a substantial part of the genetic risk for LOAD is neglected without incorporating the effects of genes other than *APOE*. Thus, although composite genetic risk indices (such as the PRS) are growing in popularity and utility, they have not been employed in the service of identifying genetic resilience for LOAD. Now, with very large numbers of LOAD samples and a more comprehensive profile of the genetic factors that confer LOAD risk, we are entering a period in which it is possible to study the interplay of genetic risk factors and genetic modifiers that reduce their penetrance.

Here, we posit the existence of common genetic variants, which have not been identified by GWAS as associated with AD as either risk or protective factors, that can help older adults remain LOAD-free despite a high genetic-risk burden. We hypothesize that there exist resilience-associated variants that lower LOAD susceptibility in a manner that is statistically independent of the effects of risk-associated alleles (or their alternative protective alleles). We tested this hypothesis by capitalizing on the most comprehensive known PRS for LOAD^18^ and *APOE* allelic status to develop two designs identifying unaffected individuals with the highest genetic likelihood of developing LOAD. Design 1 defined “resilient” individuals as normal controls with the highest PRSs for LOAD. Design 2 defined “resilient” individuals as normal controls with at least one *APOE*-_ε_4 allele and the highest LOAD PRSs (excluding the *APOE* region). We aimed to discover residual common genetic variants that confer resilience to unaffected individuals in the highest genetic risk-tiers for LOAD. We then leveraged this profile of resilience-promoting genetic variants to build a polygenic resilience score for LOAD. We hypothesized that polygenic resilience scores would account for significant variation in affection status for LOAD among individuals with high genetic risk, and would show a significant positive correlation with PRSs in unaffected controls.

## 2. Methods

### 2.1 Research design

Our workflow is shown in **Figure 1**. In stage 1, a recent GWAS meta-analysis for LOAD^18^ was leveraged for identifying risk variants and polygenic risk scoring. In stage 2, we compared two analytic designs to identify high-risk “resilient” normal controls and “risk-matched” LOAD cases. In stage 3, a resilience GWAS was conducted for each design using the identified high-risk individuals. Then the polygenic resilience score weights were derived from resilience GWAS meta-analysis summary statistics. Finally, polygenic resilience scores were replicated in independent external studies for evaluating the performance in distinguishing high-risk “resilient” normal controls from “risk-matched” LOAD cases. The parameters of each analysis step are summarized in **Supplementary Table 3**.

**Figure 1.**
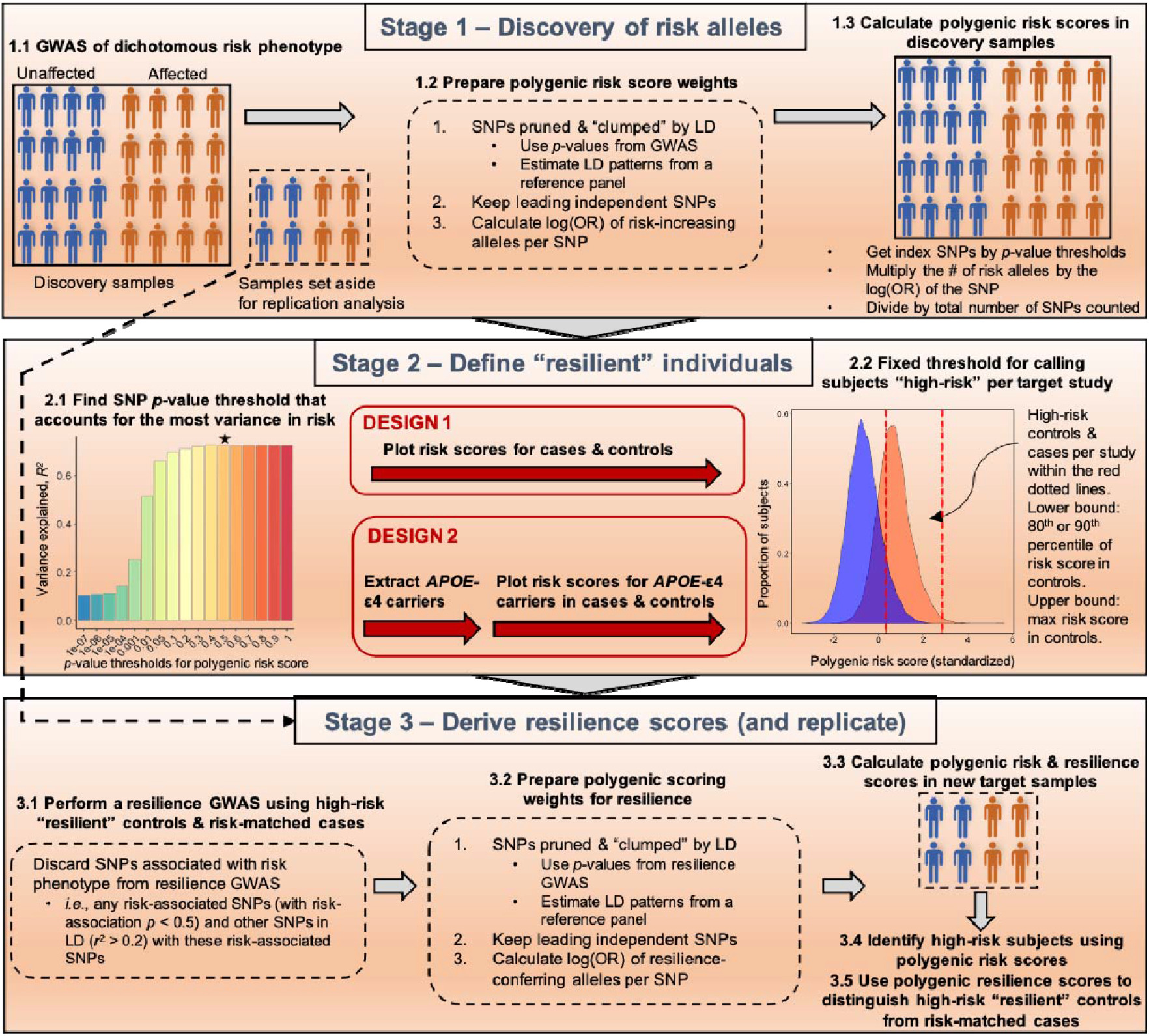
An illustration of the workflow of deriving polygenic resilience scores for late-onset Alzheimer’s disease (LOAD) for design 1 and design 2. Stage 1: Using prior LOAD GWAS results to calculate polygenic risk scores (PRSs). Stage 2: Identifying resilient individuals. In stage 2, we deployed two analysis designs differing in the definition of “resilient” individuals. In design 1, normal controls with LOAD PRSs _≥_90^th^ percentile were defined as “resilient” participants. In design 2, within the subset of normal controls who had at least one apolipoprotein E (*APOE*)-_ε_4 allele, a threshold of _≥_80^th^ percentile of PRSs (excluding SNPs in the *APOE* region) was used to define high-risk controls as “resilient”. Stage 3: Resilience genome-wide association study (GWAS) and replication of polygenic resilience scores. GWAS was performed using “resilient” individuals and risk-matched affected cases from each of the two designs. For each design, polygenic resilience scores were derived and evaluated in external replication datasets. Abbreviations: LD, linkage disequilibrium; OR, odds ratio; SNPs, single nucleotide polymorphisms.

## 2.2 Samples and genotypes

**Table 1** shows the number of normal controls, LOAD cases, high-risk “resilient” normal controls, and “risk-matched” LOAD cases in each study. Summary statistics of age-at-onset (AAO) for LOAD cases and age-at-last-examination (AAE) for normal controls are presented in **Supplementary Table 1**. In design 1 and design 2, the mean AAE of high-risk “resilient” normal controls and the mean AAO of “risk-matched” LOAD cases ranged from 70.3 to 80.9, and there were no significant age differences between groups. A common lower bound for AAO of LOAD is 65; however, the age cutoff has no specific biological significance^3^, and many genetic studies of LOAD have included cases with AAO as low as 60 (and the same AAE for unaffected comparison subjects). Therefore, we included participants in our analysis having AAO/AAE _≥_60 years old. The full name and accessibility of each study can be found in **Supplementary Table 2**. All 26 studies in the discovery stage came from the stage-1 AD GWAS meta-analysis of Kunkle *et al*.^18^. The eight studies in the replication stage are fully independent of the discovery studies. Full descriptions of the discovery and replication samples were published previously^9, 17, 18, 51, 52^. Genotypes for all studies were imputed using the Haplotype Reference Consortium (HRC) r1.1 2016 reference panel^53^. Detailed quality control (QC) steps for samples and genotypes are described in **Supplementary Methods**.

**Table 1.**
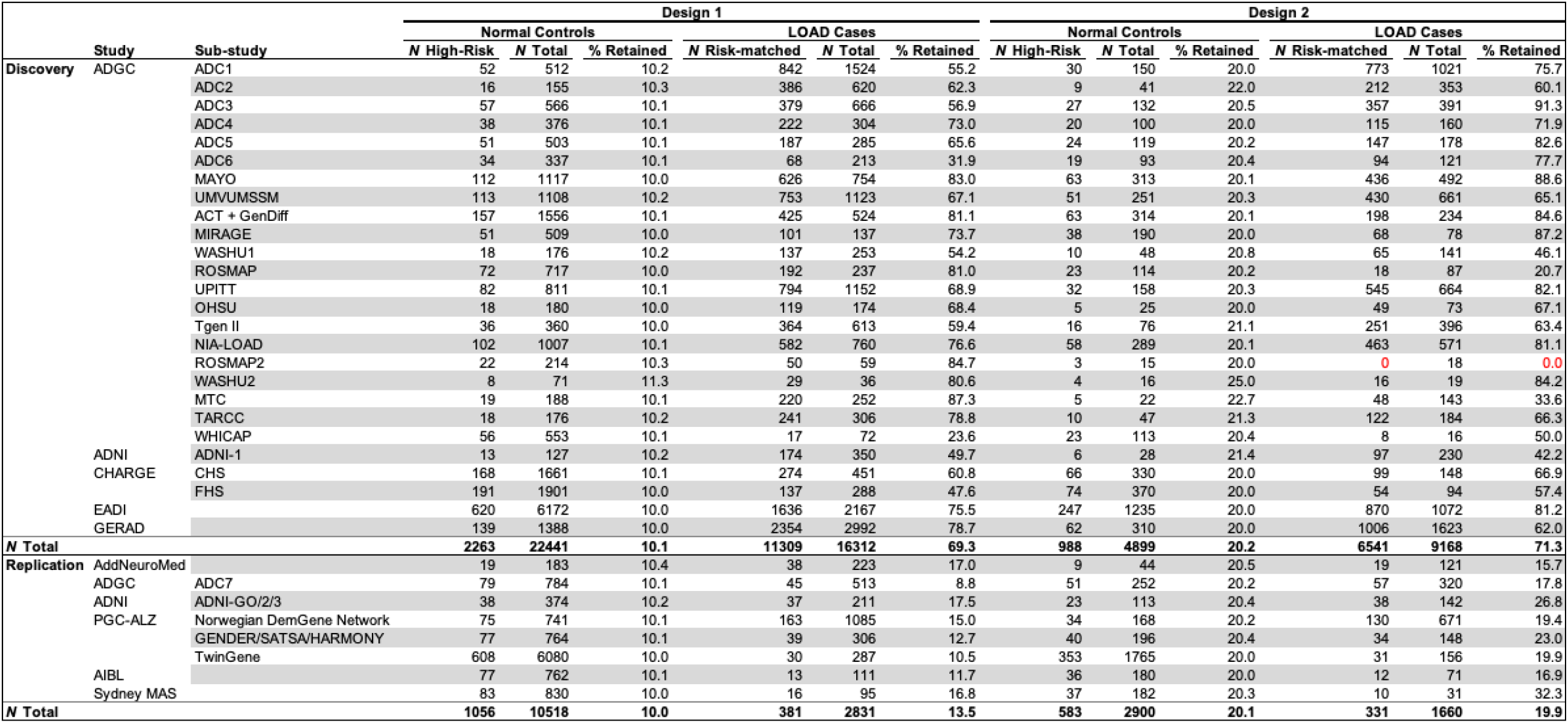
The number of LOAD cases and normal controls, high-risk normal controls (“resilient” individuals) and risk-matched LOAD cases identified in each of the discovery and replication studies. Abbreviations: LOAD, late-onset Alzheimer’s disease. A list of study full names is in Supplementary Table 2. Note: “Retained” column indicates the percentage of high-risk normal controls of all normal controls retained for resilience genome-wide association analysis per study, or the percentage of risk-matched LOAD cases of all LOAD cases retained in analysis per study. ROSMAP2 study had no LOAD cases (highlighted in red) whose risk matched with high-risk normal controls in design 2, and was not included in analysis for design 2.

### 2.3 Identifying individuals at high genetic risk

In design 1, a PRS was used to select individuals with high genetic risk. The PRS weights were derived from stage-1 AD GWAS meta-analysis summary statistics^18^. See **Supplementary Methods** for further details. The variance in AD explained by PRS maximizes at a *p*-value threshold of 0.5 in samples from GERAD (Genetic and Environmental Risk for Alzheimer’s disease)^23^, so we adopted this threshold for risk scoring. Within each study, LOAD cases and normal controls were ranked based on their PRSs. Following the original workflow^10^, the 10% of controls with the highest PRSs were classified as “resilient”. The LOAD cases whose risk scores were between the 90^th^ percentile and the maximum PRS in controls were retained as risk-matched LOAD cases for comparison.

In design 2, we restricted the analysis to *APOE*-_ε_4 carriers. *APOE* and its flanking region (chr19: 44,400kb–46,500kb)^23^ were removed from the PRS. As this analysis was restricted to fewer individuals, we lowered the high-PRS cutoff for identifying “resilient” individuals to the 80^th^ percentile to retain more participants and preserve power. In this design, “resilient” normal controls were identified as those with at least one *APOE*-_ε_4 allele, and a risk score ranked at _≥_80^th^ percentile. Risk-matched LOAD cases were defined as *APOE*-_ε_4 carriers whose PRSs fell within the high-PRS range of “resilient” normal controls.

## 2.4 Derivation, replication, and statistical analysis of polygenic resilience scores

GWASs of resilience were performed using logistic regression with *Plink* (version 1.9)^54^. Selected principal components, AAO/AAE and sex were used as covariates. A GWAS meta-analysis was conducted in *METAL*^55^ software using an inverse-variance random-effect model with genomic control. In accord with the pipeline described by Hess *et al*.^10^, SNPs known to be associated with LOAD risk were excluded from the resilience-scoring algorithm; these were defined as those SNPs that showed an association with AD risk (*p*<0.5) from the GWAS meta-analysis summary statistics^18^, and variants that were in linkage-disequilibrium (LD) (*r*^*2*^ _≥_0.2 in a 1Mb window) with those risk variants with associations of *p*<0.5. This pruning step of excluding risk variants from consideration as resilience loci serves as a conservative measure to avoid re-discovering risk variants for resilience scoring. For both resilience designs, the polygenic resilience score weights were generated from the marginal SNPs of resilience GWAS meta-analysis summary statistics following the same series of QC steps (see **Supplementary Methods**).

Polygenic resilience scores were derived for 10 *p*-value thresholds, in a manner similar to the PRS algorithm, by summing up the weighted effective allele counts of SNPs^26^. Logistic regression was used to assess the likelihood of “resilient” group inclusion based on harboring a higher polygenic resilience score. Selected principal components, AAO/AAE and sex were used as covariates. For each polygenic resilience score, we meta-analyzed the natural logarithm of the odds ratio (OR) of being a high-risk “resilient” normal control versus a risk-matched LOAD case using a random-effects inverse-variance model using the *R* package *metafor*, and pooled variance explained in resilience across independent replication studies. All tests were two-tailed. See **Supplementary Methods** for further details.

## 3. Results

### 3.1 Resilience GWAS

Design 1 produced 2,263 high-risk “resilient” normal controls and 11,309 risk-matched LOAD cases for the resilience GWAS meta-analysis. As expected, the sample size retained in design 2 was smaller, totaling 988 high-risk “resilient” normal controls and 6,541 risk-matched LOAD cases (**Table 1**). Because our analytic approaches used only subsets of all available LOAD case-control GWAS data, we neither had nor anticipated having sufficient power to detect individual SNPs with genome-wide significant association with resilience (**Supplementary Figure 3**). Instead, our focus was on deriving and evaluating polygenic resilience scores. As a necessary step to generate SNP-weights for summation in those scores, we performed individual-SNP association tests and briefly reported the results in **Supplementary Results**.

### 3.2 Replication and evaluation of polygenic resilience scores

After removing risk-associated SNPs (*p*<0.5) and SNPs in LD with those risk-associated SNPs (*r*^*2*^ _≥_0.2), clumping the remaining marginal SNPs, and applying QC steps, a profile of 18,723 SNPs was included in the resilience score for design 1, and 18,122 SNPs in design 2. Resilience scores for all 10 *p*-value thresholds were significantly associated with “resilient” group inclusion (“resilient” normal controls versus risk-matched LOAD cases) when tested in locally downloaded discovery datasets. Results of the association between “resilient” group inclusion and polygenic resilience scores from the replication datasets were meta-analyzed, yielding 1,056 high-risk “resilient” normal controls and 381 risk-matched LOAD cases in design 1, and 583 high-risk “resilient” normal controls and 331 risk-matched LOAD cases in design 2 (**Table 1**).

In design 1, the meta-analysis found significant replication of the association between “resilient” group inclusion and polygenic resilience scores at two *p*-value thresholds (*p*<0.1, *p*<0.2) (**Figure 2A**). The most significant association was found for the polygenic resilience score containing all independent marginal SNPs with resilience GWAS *p*<0.1 (OR=1.24, 95% confidence interval [CI]=1.05-1.47, *p*=0.010). Resilience scores for the 0.1 *p*-value threshold explained an average of 1.3% (standard deviation [SD]=5.3%) of the variance in “resilient” group inclusion or 1.2% (SD=4.3%) (**Figure 2B**) of the variance on the liability scale, *i*.*e*., SNP-heritability of resilience. No significant (*p*<0.05) replication of the association between “resilient” group inclusion and polygenic resilience scores was observed for any of the 10 polygenic resilience scores in design 2.

**Figure 2.**
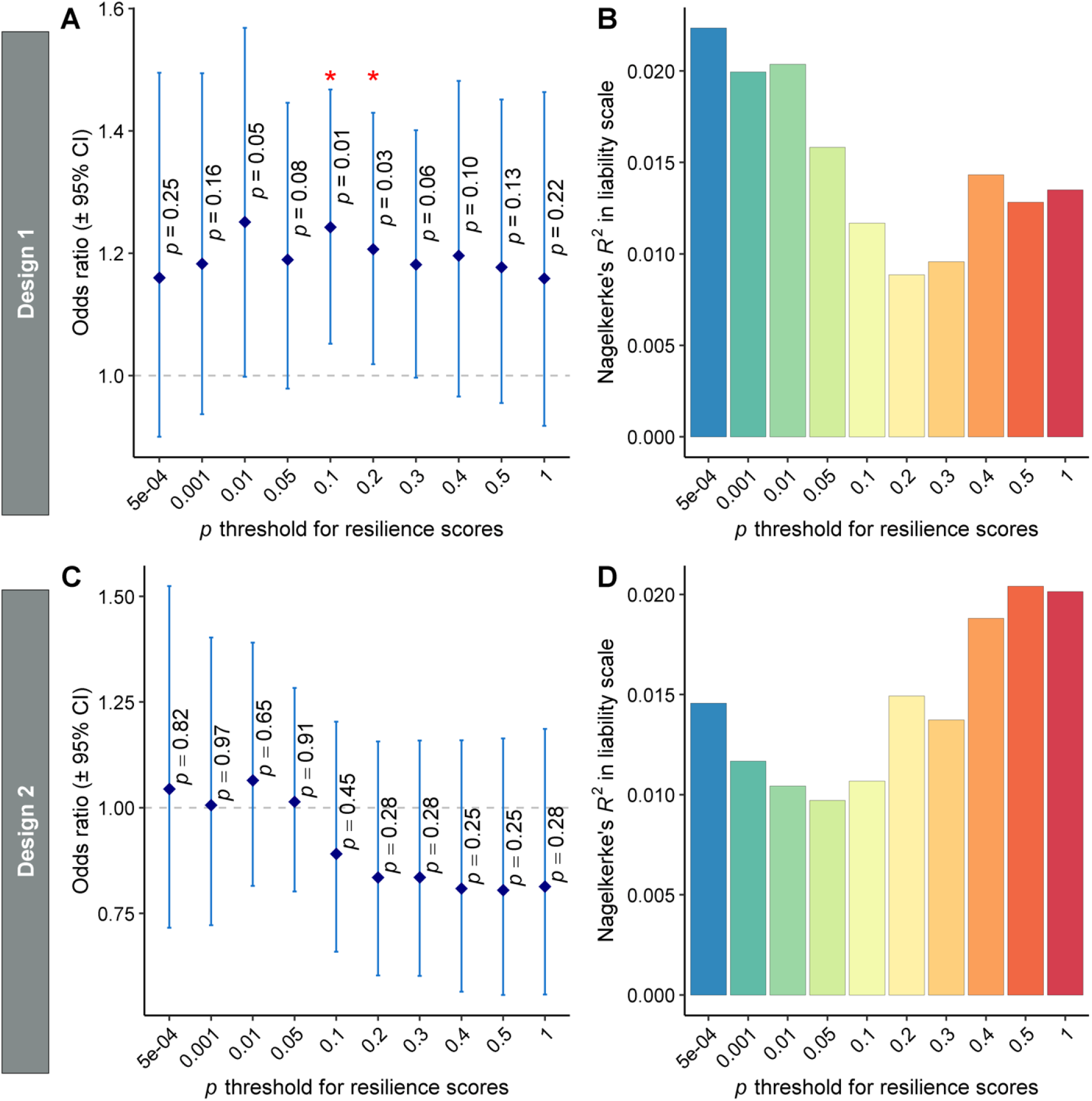
The performance of polygenic resilience scores in capturing resilience variability in independent replication studies. In design 1, normal controls with late-onset Alzheimer’s disease (LOAD) polygenic risk scores (PRSs) _≥_90^th^ percentile were defined as “resilient” participants. In design 2, a threshold of _≥_80^th^ percentile of PRSs (excluding SNPs in the apolipoprotein E [*APOE*] region) was used to define high-risk controls as “resilient” within the normal controls who have at least one *APOE*-_ε_4 allele. Panels A and B: design 1 (high-risk normal controls, *n* = 1,056; risk-matched LOAD cases, *n* = 381). Panels C and D: design 2 (high-risk normal controls, *n* = 583; risk-matched LOAD cases, *n* = 331). The odds ratio (OR) and variance explained by polygenic resilience scores reflect meta-analytic results from independent replication samples. Nagelkerke’s pseudo-*R*^2^ values in the liability scale are weighted average using the weights from the meta-analysis of ORs. The dot-plots (Panels A and C) show corresponding ORs for resilience scores across 10 *p*-value thresholds, wherein OR > 1.0 indicates higher resilience scores are associated with a higher likelihood of being a high-risk normal control (“resilient” individual) than being a risk-matched LOAD case. Error bars represent the 95% confidence intervals (CI) around each OR, which are the exponent of the 95% CI of β coefficients. The bar-plots (Panels B and D) show the amount of variance in resilience (*i*.*e*., “resilient” high-risk normal controls versus risk-matched LOAD cases) on the liability scale that is explained by resilience scores. Asterisks (*) indicate *p*-values < 0.05 for ORs > 1.0.

### 3.3 Interaction of risk and resilience effects

In the full samples from three locally downloaded replication studies (Alzheimer Disease Centers Wave 7 [ADC7], AddNeuroMed, and Alzheimer’s Disease Neuroimaging Initiative stage GO/2/3 [ADNI-GO/2/3]; normal controls, *n*=1,321; LOAD cases, *n*=943) (**Table 1** and **Supplementary Table 1**), we tested for correlations between PRSs and polygenic resilience scores. As hypothesized, the standardized polygenic resilience scores of the optimal *p*<0.1 threshold in design 1 exhibited a significant positive correlation with PRSs in normal controls (Pearson’s *r*=0.102, 95% CI=0.048-0.155, degree of freedom [*df*]=1319, *p*=2.1e-04), and no significant correlation was observed in LOAD cases (Pearson’s *r*=0.022, 95% CI=-0.042-0.085, *df* =941, *p*=0.51) (**Figure 3**).

**Figure 3.**
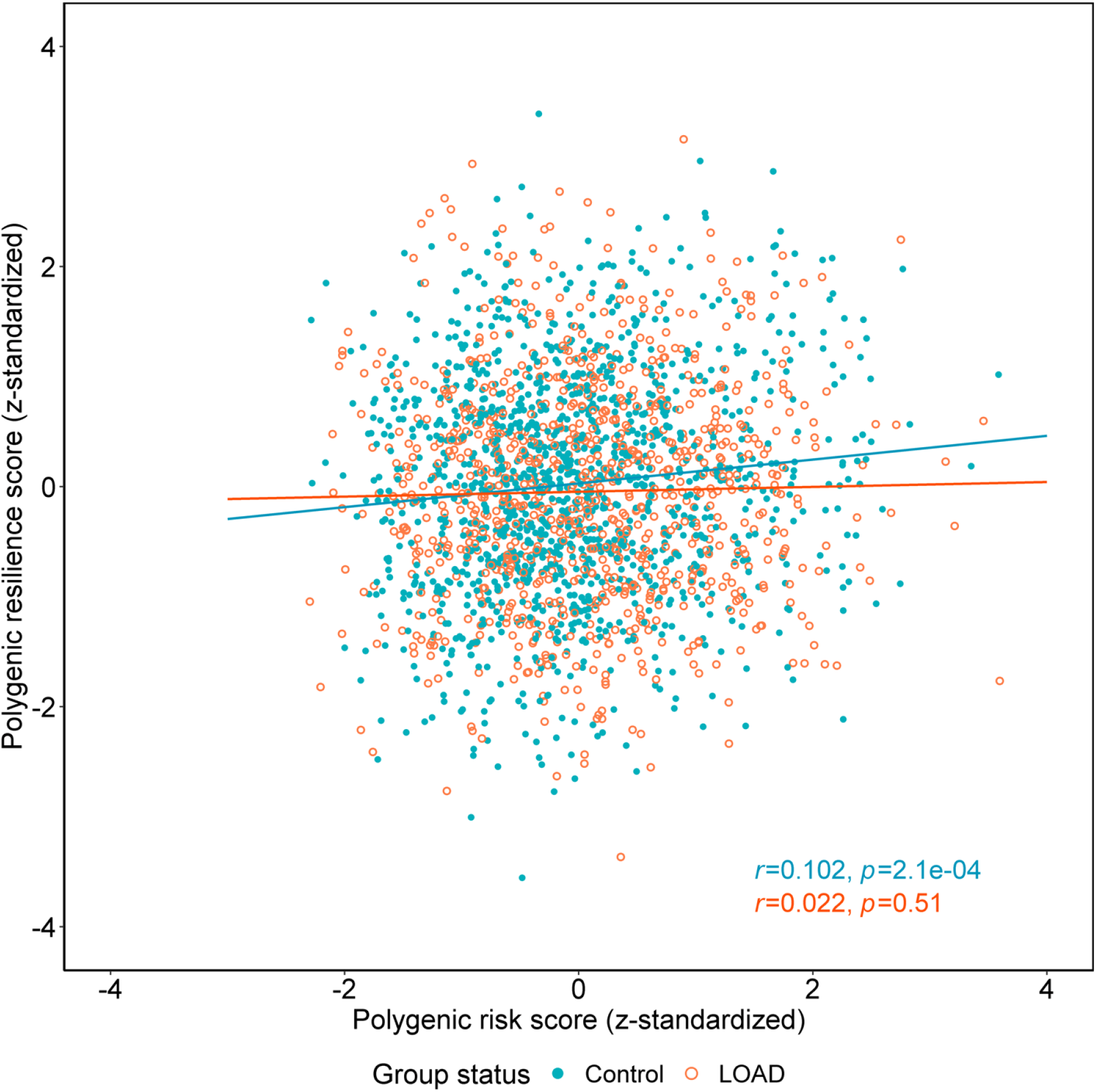
**The correlation of standardized polygenic risk scores (PRSs) and polygenic resilience scores (design 1) in normal controls and late-onset Alzheimer’s disease (LOAD) cases** in three independent replication studies not used in the resilience score derivation steps (*i*.*e*., ADC7, AddNeuroMed, and ADNI-GO/2/3; normal controls, *n* = 1,321; LOAD cases, *n* = 943). The optimal *p*-value threshold for polygenic risk-scoring was 0.5, and the optimal *p-*value threshold for polygenic resilience scoring was 0.1 (see Figure 2). The blue round dots indicate normal controls, and the orange circles indicate LOAD cases. The blue and orange lines represent the best-fit for correlations between PRSs and resilience scores in normal controls and in LOAD cases, respectively. The blue and orange annotation text shows the Pearson correlation coefficient (*r*) and the *p*-value between PRSs and resilience scores in normal controls and LOAD cases, respectively. In this analysis, we excluded ultra-high-risk LOAD cases whose PRSs are higher than the maximum of all normal controls, and ultra-low-risk normal controls whose PRSs are lower than the minimum of all LOAD cases.

## 4. Discussion

We applied a validated analytic framework to detect common variants that, when combined into a polygenic resilience score, are associated with lower LOAD-risk penetrance among older individuals with relatively high genetic risk of disease. We found reliable evidence to reinforce the notion that unaffected individuals with higher genetic risk-loads may be protected from complex diseases, such as LOAD, by the collective effects of risk-independent common variants that reduce the penetrance of one’s overall genetic risk burden. Identifying genetic factors that moderate risk penetrance may prove valuable for explaining the missing heritability and etiologic heterogeneity of LOAD, which in turn could shed light on pathophysiological mechanisms and eventually lead to better interventions and preventive treatments.

### 4.1 Risk-countering effects of polygenic resilience scores

Individuals with higher polygenic resilience scores (*p*<0.1 and *p*<0.2 thresholds of design 1) had higher odds of being a “resilient” high-risk normal control than a risk-matched LOAD case. Polygenic resilience scores (design 1) significantly increased with higher PRSs in normal controls, but not in LOAD cases. Taken together, these results support the hypothesis that polygenic resilience scores capture risk-countering polygenic effects against the penetrance of high polygenic risk for LOAD, and that normal controls with higher PRSs are protected from LOAD by harboring correspondingly higher polygenic resilience scores. Although no polygenic resilience scores in design 2 demonstrated significant risk-buffering effects, we cannot rule out the possibility that common variants might reduce risk penetrance in normal controls with enriched risk from both *APOE* and PRSs. In fact, among *APOE*-_ε_4 carriers, higher resilience scores in design 1 at the *p*<0.1 threshold (OR=1.64, 95% CI=1.08-2.50, *p*=0.021) and the *p*<0.2 threshold (OR=1.98, 95% CI=1.24-3.15, *p*=3.9e-03) were associated with higher odds of being a “resilient” high-PRS normal control than a risk-matched LOAD case. Among “resilient” high-PRS controls, higher resilience scores in design 1 (*p*<0.2 threshold) were significantly associated with increased odds of carrying at least one *APOE*-_ε_4 allele (OR=1.57, 95% CI=1.07-2.29, *p*=0.021). A similar trend was observed when the *p*<0.1 threshold was used, although this was not significant (OR=1.30, 95% CI=0.90-1.89, *p*=0.16) (**Supplementary Results**). We therefore conclude that polygenic resilience scores may moderate the risk effects of the LOAD PRS generally, and the *APOE*-_ε_4 allele specifically. However, these analyses were carried out in relatively small studies (ADC7, AddNeuroMed, and ADNI-GO/2/3), and need to be repeated in larger, more powerful, replication samples.

### 4.2 Interplay of polygenic effects and APOE

In design 2, we hypothesized that a two-stage selection of individuals (with both higher PRSs and one or more *APOE*-_ε_4 alleles) would enrich for individuals with the absolute highest genetic risk for LOAD^56, 57^; yet, there was a substantial reduction in the performance of design 2 in contrast to design 1. The lack of significant replication of association with resilience in design 2 simply might be due to lower statistical power in both the resilience score development and replication stages, considering the total sample size of design 2 is approximately half that of design 1. Alternatively, resilience-promoting variants may be found among *APOE*-_ε_4 carriers through broader exploration of the model-parameter space (*e*.*g*., PRS threshold in particular), separate evaluation of *APOE*-_ε_4 homozygotes and various heterozygote combinations, and more accurate modeling of the genetic architecture of resilience (see limitations below). An important question future studies should address is to what extent common variants may influence the penetrance of genetic risk in larger samples of *APOE*-_ε_4 carriers, or whether the prevalence of risk-modifying common variants differs between *APOE*-_ε_4 carriers and non-carriers.

On the other hand, multiple studies^22-25, 28, 58-61^ have revealed that PRSs capture independent risk effects beyond *APOE* alone, while few studies have explored the risk-predictive performance of PRSs stratified by *APOE* status. Higher PRSs were found to be associated with increased susceptibility for LOAD in *APOE-*_ε_4 non-carriers^25, 29, 56^. Furthermore, the risk effects of PRS deciles across *APOE* status could be dependent on the ages of participants^29, 56, 60, 62^. Further mining of the complex relationship between the risk effects of PRSs and *APOE* is outside the scope of the current study; however, further investigations on the penetrance of high PRSs among *APOE*-_ε_4 carriers and non-carriers seem warranted.

### 4.3 Strengths and limitations

Our approach has identified candidate resilience loci that may ultimately serve as targets for the promotion of resilience. We examined the performance of two polygenic resilience scores: design 1 selected participants with the highest polygenic risk regardless of *APOE*-_ε_4 status, while design 2 restricted analyses to *APOE*-_ε_4 carriers. To our knowledge, this is the first study to identify a polygenic resilience score for genetic LOAD risk, comprising thousands of risk-independent common variants that partially offset the genetic risk conferred by a relatively high PRS. An important distinction of the current study relative to prior work on genetic resilience to LOAD is that we accounted not only for the risk from *APOE*, but also the aggregate effect of thousands of additional risk variants throughout the genome *via* the LOAD PRS.

A conservative variant-filtering strategy was applied, which resulted in the removal of common variants associated with LOAD risk variants (risk association *p*<0.5) and those in liberal LD (*r*^*2*^>0.2) with LOAD risk variants. A strength of this approach is that we ensured the polygenic resilience scores derived in the current study are independent of the risk scores so that the SNPs comprising the polygenic resilience score are not sub-threshold risk SNPs. Additionally, the resilience alleles of these risk-residual SNPs are not simply protective alleles defined in a risk framework, where each biallelic locus is defined by both a risk allele and a corresponding and opposing protective allele. Thus, this strategy helps identify resilience effects that are conditioned on net risk effects, owing to the combination of risk and protective alleles summed in polygenic risk scores. Yet, although our approach is conservative, it is limited in the identification of a better-performing resilience score because most of the genome has been discarded from the analysis. Biologically, it is plausible that variants nearby risk loci, such as those in the same LD block or in the same gene with risk SNPs, could exert modifying functions^63^. Our conservative strategy, discarding all SNPs with any semblance of risk-association, and those in liberal LD threshold with such SNPs, consequently leads to lower power in uncovering variants with potentially higher biological functionality. This notion is borne out in the fact that no significant gene-ontology pathways were enriched by resilience-related common variants identified in this study (results not shown). With larger samples, resilience-conferring SNPs may be investigated using a stricter LD threshold (*e*.*g*., *r*^*2*^>0.1) to further restrain the “hitchhiking” of risk variants. More importantly, Mendelian randomization, conditional association testing, or simulation analysis may be better suited to evaluate the hypothesis that resilience signals are more likely to co-localize with risk loci or genes. In addition, filtering variants by LD with risk SNPs results in a low LD structure among the remaining SNPs as demonstrated previously^10^, which diminishes our capacity to examine the genetic correlation of resilience to LOAD with other risk- or resilience-related phenotypes (*e*.*g*., *via* LD score regression). A high priority should be placed on the design of new methods that can detect resilience-associated SNPs that may reside in regions of strong LD with risk variants.

Resilience was defined by discrete groups in our analysis, which truncated effective sample sizes to the upper tail of the risk distribution. In the future, when larger samples are available, higher risk thresholds can be applied and more extreme-risk samples could be leveraged to increase statistical power. Theoretically, resilience also could be a continuous measure. Thus, our resilience approach might also be improved by leveraging all study samples and modeling the continuity of resilience using either linear or non-linear analysis. Despite the restricted sample sizes in the current study, two *p*-value thresholds of the resilience scores in design 1 were sufficiently robust to replicate significantly in fully independent studies. Further replication would be key to testing the validity of these resilience scores. It is expected that the strength of our results (in terms of variance-explained and the significance of associations) will only increase with the addition of more samples.

Several studies^9, 22, 58, 64^ indicated that polygenic risk scores of *p*-value thresholds less than 0.5 (*i*.*e*., 5*e*^-08^, 1*e*^-05^) might show better performance in predicting LOAD risk. Therefore, it may be valuable to compare the performance of resilience scores developed from risk scores at other *p*-value thresholds. In addition, it is likely that a subgroup of “resilient” normal controls identified in the present work will eventually develop LOAD, but with later onset. Thus, all resilient participants demonstrate resilience against high levels of genetic risk for LOAD, but only those who never develop LOAD are additionally resistant against the disease itself. Lastly, the participants in our analysis were of European ancestry, so the degree of generalization of our results to non-European populations is presently unknown.

### 4.4 Future directions

Two analysis designs were deployed in the current study to select individuals with a high genetic risk burden from both PRSs and *APOE*, and other methods could be devised to expand the capabilities of our resilience approach in LOAD. It has been suggested, for example, that using a PRS with the *APOE* region removed and adding *APOE* alleles as a covariate may boost the performance of LOAD risk prediction^65^, compared with incorporating *APOE* alleles as weighted SNPs in PRSs. In addition, it could be important to include the number of *APOE*-_ε_4 or _ε_2 alleles as covariates in resilience analysis models to better reflect the relative risk levels among individuals.

Potentially, polygenic resilience scores from the current study could be applied to other resilience-related questions. For example, it would be instrumental in discovering the extent to which polygenic resilience score is associated with other phenotypes that have been associated with resilience to LOAD risk (*e*.*g*., education, general cognitive ability in early life, and other indices of cognitive reserve, brain reserve, or brain maintenance)^31, 32, 66-70^. In follow-up studies, it might be illuminating to investigate whether these resilience-promoting genetic factors show protective effects for cognitive impairment or LOAD-related pathophysiological changes.

## 4.5 Conclusion

We found evidence to support the hypothesis that thousands of risk-independent common variants underlie resilience among unaffected individuals with higher genetic risk for LOAD. We conclude that common variants not in LD with known LOAD risk variants exert a protective effect on LOAD risk. Our findings provide a significant and novel contribution to the existing understanding of genetic resilience to LOAD risk. This novel approach highlights a window of opportunity for identifying risk-modifying biological mechanisms and potential pathways for intervention in populations at the highest risk for LOAD.

## Data Availability

All data produced in the present study are available upon reasonable request to the authors

## Data availability

Resilience-scoring weights for design 1 can be shared by request. The Alzheimer’s Disease Genetics Consortium (ADGC), the Alzheimer’s Disease Neuroimaging Initiative (ADNI), and the AddNeuroMed data used in this study were provided under restricted access by NIAGADS (https://www.niagads.org), ADNI (http://adni.loni.usc.edu), and Synapse platform (https://www.synapse.org/#!Synapse:syn4907804), respectively. Only summary statistics were made available to us from the European Alzheimer’s Disease Initiative (EADI), the Genetic and Environmental Risk in Alzheimer’s Disease (GERAD), the Cohorts for Heart and Aging Research in Genomic Epidemiology (CHARGE), the Psychiatric Genomic Consortium (PGC), the Australian Imaging, Biomarker & Lifestyle Study (AIBL), and the Sydney Memory and Ageing Study (Sydney MAS).

## Acknowledgements

We thank all the participants of this study for their contributions. Our effort on this project was supported by the U.S. National Institute on Aging [grant numbers R01AG064955 and R01AG054002]. Additional acknowledgements and detailed acknowledgements of funding sources for the study are provided in **Supplementary acknowledgements**.

## Abbreviations

AD: Alzheimer’s disease
LOAD: late-onset Alzheimer’s disease
SNPs: single nucleotide polymorphisms
GWASs: genome-wide association studies
*APOE*: apolipoprotein E
PRS: polygenic risk score
AAO: age-at-onset
AAE: age-at-last-examination
QC: quality-control
HRC: Haplotype Reference Consortium
GERAD: Genetic and Environmental Risk for Alzheimer’s disease
LD: linkage-disequilibrium
OR: odds ratio
CI: confidence interval
SD: standard deviation
df: degree of freedom
ADNI: Alzheimer’s Disease Neuroimaging Initiative
ADC7: Alzheimer Disease Centers Wave 7

## References

1. 2021 Alzheimer’s disease facts and figures. Alzheimers Dement 2021; 17(3): 327–406.

2. Ryan NS, Nicholas JM, Weston PSJ, Liang Y, Lashley T, Guerreiro R et al. Clinical phenotype and genetic associations in autosomal dominant familial Alzheimer’s disease: a case series. Lancet Neurol 2016; 15(13): 1326–1335.

3. Rossor MN, Fox NC, Mummery CJ, Schott JM, Warren JD. The diagnosis of young-onset dementia. Lancet Neurol 2010; 9(8): 793–806.

4. Edwards Iii GA, Gamez N, Escobedo G, Jr., Calderon O, Moreno-Gonzalez I. Modifiable Risk Factors for Alzheimer’s Disease. Front Aging Neurosci 2019; 11: 146.

5. Gatz M, Reynolds CA, Fratiglioni L, Johansson B, Mortimer JA, Berg S et al. Role of genes and environments for explaining Alzheimer disease. Arch Gen Psychiatry 2006; 63(2): 168–174.

6. Lee SH, Harold D, Nyholt DR, Consortium AN, International Endogene C, Genetic et al. Estimation and partitioning of polygenic variation captured by common SNPs for Alzheimer’s disease, multiple sclerosis and endometriosis. Hum Mol Genet 2013; 22(4): 832–841.

7. Ridge PG, Mukherjee S, Crane PK, Kauwe JS, Alzheimer’s Disease Genetics C. Alzheimer’s disease: analyzing the missing heritability. PLoS One 2013; 8(11): e79771.

8. Brainstorm C, Anttila V, Bulik-Sullivan B, Finucane HK, Walters RK, Bras J et al. Analysis of shared heritability in common disorders of the brain. Science 2018; 360(6395).

9. Zhang Q, Sidorenko J, Couvy-Duchesne B, Marioni RE, Wright MJ, Goate AM et al. Risk prediction of late-onset Alzheimer’s disease implies an oligogenic architecture. Nat Commun 2020; 11(1): 4799.

10. Hess JL, Tylee DS, Mattheisen M, Schizophrenia Working Group of the Psychiatric Genomics C, Lundbeck Foundation Initiative for Integrative Psychiatric R, Borglum AD et al. A polygenic resilience score moderates the genetic risk for schizophrenia. Mol Psychiatry 2021; 26(3): 800–815.

11. Harold D, Abraham R, Hollingworth P, Sims R, Gerrish A, Hamshere ML et al. Genome-wide association study identifies variants at CLU and PICALM associated with Alzheimer’s disease. Nat Genet 2009; 41(10): 1088–1093.

12. Lambert JC, Heath S, Even G, Campion D, Sleegers K, Hiltunen M et al. Genome-wide association study identifies variants at CLU and CR1 associated with Alzheimer’s disease. Nat Genet 2009; 41(10): 1094–1099.

13. Hollingworth P, Harold D, Sims R, Gerrish A, Lambert JC, Carrasquillo MM et al. Common variants at ABCA7, MS4A6A/MS4A4E, EPHA1, CD33 and CD2AP are associated with Alzheimer’s disease. Nat Genet 2011; 43(5): 429–435.

14. Naj AC, Jun G, Beecham GW, Wang LS, Vardarajan BN, Buros J et al. Common variants at MS4A4/MS4A6E, CD2AP, CD33 and EPHA1 are associated with late-onset Alzheimer’s disease. Nat Genet 2011; 43(5): 436–441.

15. Lambert JC, Ibrahim-Verbaas CA, Harold D, Naj AC, Sims R, Bellenguez C et al. Meta-analysis of 74,046 individuals identifies 11 new susceptibility loci for Alzheimer’s disease. Nat Genet 2013; 45(12): 1452–1458.

16. Marioni RE, Harris SE, Zhang Q, McRae AF, Hagenaars SP, Hill WD et al. GWAS on family history of Alzheimer’s disease. Transl Psychiatry 2018; 8(1): 99.

17. Jansen IE, Savage JE, Watanabe K, Bryois J, Williams DM, Steinberg S et al. Genome-wide meta-analysis identifies new loci and functional pathways influencing Alzheimer’s disease risk. Nat Genet 2019; 51(3): 404–413.

18. Kunkle BW, Grenier-Boley B, Sims R, Bis JC, Damotte V, Naj AC et al. Genetic meta-analysis of diagnosed Alzheimer’s disease identifies new risk loci and implicates Abeta, tau, immunity and lipid processing. Nat Genet 2019; 51(3): 414–430.

19. Wightman DP, Jansen IE, Savage JE, Shadrin AA, Bahrami S, Rongve A et al. Largest GWAS (N=1,126,563) of Alzheimer’s Disease Implicates Microglia and Immune Cells. medRxiv 2020.

20. Schwartzentruber J, Cooper S, Liu JZ, Barrio-Hernandez I, Bello E, Kumasaka N et al. Genome-wide meta-analysis, fine-mapping and integrative prioritization implicate new Alzheimer’s disease risk genes. Nat Genet 2021; 53(3): 392–402.

21. Corder EH, Saunders AM, Strittmatter WJ, Schmechel DE, Gaskell PC, Small GW et al. Gene dose of apolipoprotein E type 4 allele and the risk of Alzheimer’s disease in late onset families. Science 1993; 261(5123): 921–923.

22. Altmann A, Scelsi MA, Shoai M, de Silva E, Aksman LM, Cash DM et al. A comprehensive analysis of methods for assessing polygenic burden on Alzheimer’s disease pathology and risk beyond APOE. Brain Commun 2020; 2(1): fcz047.

23. Escott-Price V, Sims R, Bannister C, Harold D, Vronskaya M, Majounie E et al. Common polygenic variation enhances risk prediction for Alzheimer’s disease. Brain 2015; 138(Pt 12): 3673–3684.

24. Escott-Price V, Shoai M, Pither R, Williams J, Hardy J. Polygenic score prediction captures nearly all common genetic risk for Alzheimer’s disease. Neurobiol Aging 2017; 49: 214 e217–214 e211.

25. Escott-Price V, Myers A, Huentelman M, Shoai M, Hardy J. Polygenic risk score analysis of Alzheimer’s disease in cases without APOE4 or APOE2 alleles. J Prev Alzheimers Dis 2019; 6(1): 16–19.

26. Schizophrenia Working Group of the Psychiatric Genomics C. Biological insights from 108 schizophrenia-associated genetic loci. Nature 2014; 511(7510): 421–427.

27. Karlsson IK, Escott-Price V, Gatz M, Hardy J, Pedersen NL, Shoai M et al. Measuring heritable contributions to Alzheimer’s disease: polygenic risk score analysis with twins. Brain communications 2022; 4(1): fcab308.

28. Ge T, Sabuncu MR, Smoller JW, Sperling RA, Mormino EC, Alzheimer’s Disease Neuroimaging I. Dissociable influences of APOE epsilon4 and polygenic risk of AD dementia on amyloid and cognition. Neurology 2018; 90(18): e1605–e1612.

29. Najar J, van der Lee SJ, Joas E, Wetterberg H, Hardy J, Guerreiro R et al. Polygenic risk scores for Alzheimer’s disease are related to dementia risk in APOE varepsilon4 negatives. Alzheimers Dement (Amst) 2021; 13(1): e12142.

30. Khachaturian ZS, Petersen RC, Snyder PJ, Khachaturian AS, Aisen P, de Leon M et al. Developing a global strategy to prevent Alzheimer’s disease: Leon Thal Symposium 2010. Alzheimers Dement 2011; 7(2): 127–132.

31. Stern Y, Arenaza-Urquijo EM, Bartres-Faz D, Belleville S, Cantilon M, Chetelat G et al. Whitepaper: Defining and investigating cognitive reserve, brain reserve, and brain maintenance. Alzheimers Dement 2020; 16(9): 1305–1311.

32. Perneczky R, Kempermann G, Korczyn AD, Matthews FE, Ikram MA, Scarmeas N et al. Translational research on reserve against neurodegenerative disease: consensus report of the International Conference on Cognitive Reserve in the Dementias and the Alzheimer’s Association Reserve, Resilience and Protective Factors Professional Interest Area working groups. BMC Med 2019; 17(1): 47.

33. Latimer CS, Burke BT, Liachko NF, Currey HN, Kilgore MD, Gibbons LE et al. Resistance and resilience to Alzheimer’s disease pathology are associated with reduced cortical pTau and absence of limbic-predominant age-related TDP-43 encephalopathy in a community-based cohort. Acta Neuropathol Commun 2019; 7(1): 91.

34. Arenaza-Urquijo EM, Vemuri P. Improving the resistance and resilience framework for aging and dementia studies. Alzheimers Res Ther 2020; 12(1): 41.

35. Choi KW, Stein MB, Dunn EC, Koenen KC, Smoller JW. Genomics and psychological resilience: a research agenda. Mol Psychiatry 2019; 24(12): 1770–1778.

36. Livingston G, Huntley J, Sommerlad A, Ames D, Ballard C, Banerjee S et al. Dementia prevention, intervention, and care: 2020 report of the Lancet Commission. Lancet 2020; 396(10248): 413–446.

37. Livingston G, Sommerlad A, Orgeta V, Costafreda SG, Huntley J, Ames D et al. Dementia prevention, intervention, and care. Lancet 2017; 390(10113): 2673–2734.

38. Kremen WS, Beck A, Elman JA, Gustavson DE, Reynolds CA, Tu XM et al. Influence of young adult cognitive ability and additional education on later-life cognition. Proc Natl Acad Sci U S A 2019; 116(6): 2021–2026.

39. Lee JJ, Wedow R, Okbay A, Kong E, Maghzian O, Zacher M et al. Gene discovery and polygenic prediction from a genome-wide association study of educational attainment in 1.1 million individuals. Nat Genet 2018; 50(8): 1112–1121.

40. Polderman TJ, Benyamin B, de Leeuw CA, Sullivan PF, van Bochoven A, Visscher PM et al. Meta-analysis of the heritability of human traits based on fifty years of twin studies. Nat Genet 2015; 47(7): 702–709.

41. Davies G, Lam M, Harris SE, Trampush JW, Luciano M, Hill WD et al. Study of 300,486 individuals identifies 148 independent genetic loci influencing general cognitive function. Nat Commun 2018; 9(1): 2098.

42. Farrer LA, Cupples LA, Haines JL, Hyman B, Kukull WA, Mayeux R et al. Effects of age, sex, and ethnicity on the association between apolipoprotein E genotype and Alzheimer disease. A meta-analysis. APOE and Alzheimer Disease Meta Analysis Consortium. JAMA 1997; 278(16): 1349–1356.

43. Jonsson T, Atwal JK, Steinberg S, Snaedal J, Jonsson PV, Bjornsson S et al. A mutation in APP protects against Alzheimer’s disease and age-related cognitive decline. Nature 2012; 488(7409): 96–99.

44. DeTure MA, Dickson DW. The neuropathological diagnosis of Alzheimer’s disease. Mol Neurodegener 2019; 14(1): 32.

45. Reed BR, Mungas D, Farias ST, Harvey D, Beckett L, Widaman K et al. Measuring cognitive reserve based on the decomposition of episodic memory variance. Brain 2010; 133(Pt 8): 2196–2209.

46. Negash S, Bennett DA, Wilson RS, Schneider JA, Arnold SE. Cognition and neuropathology in aging: multidimensional perspectives from the Rush Religious Orders Study and Rush Memory And Aging Project. Curr Alzheimer Res 2011; 8(4): 336–340.

47. Hohman TJ, McLaren DG, Mormino EC, Gifford KA, Libon DJ, Jefferson AL et al. Asymptomatic Alzheimer disease: Defining resilience. Neurology 2016; 87(23): 2443–2450.

48. Ayers KL, Mirshahi UL, Wardeh AH, Murray MF, Hao K, Glicksberg BS et al. A loss of function variant in CASP7 protects against Alzheimer’s disease in homozygous APOE epsilon4 allele carriers. BMC Genomics 2016; 17 Suppl 2: 445.

49. DeKosky ST, Aston CE, Kamboh MI. Polygenic determinants of Alzheimer’s disease: modulation of the risk by alpha-1-antichymotrypsin. Ann N Y Acad Sci 1996; 802: 27–34.

50. Kamboh MI, Sanghera DK, Ferrell RE, DeKosky ST. APOE*4-associated Alzheimer’s disease risk is modified by alpha 1-antichymotrypsin polymorphism. Nat Genet 1995; 10(4): 486–488.

51. Proitsi P, Lupton MK, Velayudhan L, Newhouse S, Fogh I, Tsolaki M et al. Genetic predisposition to increased blood cholesterol and triglyceride lipid levels and risk of Alzheimer disease: a Mendelian randomization analysis. PLoS Med 2014; 11(9): e1001713.

52. Lovestone S, Francis P, Kloszewska I, Mecocci P, Simmons A, Soininen H et al. AddNeuroMed--the European collaboration for the discovery of novel biomarkers for Alzheimer’s disease. Ann N Y Acad Sci 2009; 1180: 36–46.

53. McCarthy S, Das S, Kretzschmar W, Delaneau O, Wood AR, Teumer A et al. A reference panel of 64,976 haplotypes for genotype imputation. Nat Genet 2016; 48(10): 1279–1283.

54. Chang CC, Chow CC, Tellier LC, Vattikuti S, Purcell SM, Lee JJ. Second-generation PLINK: rising to the challenge of larger and richer datasets. Gigascience 2015; 4: 7.

55. Willer CJ, Li Y, Abecasis GR. METAL: fast and efficient meta-analysis of genomewide association scans. Bioinformatics 2010; 26(17): 2190–2191.

56. van der Lee SJ, Wolters FJ, Ikram MK, Hofman A, Ikram MA, Amin N et al. The effect of APOE and other common genetic variants on the onset of Alzheimer’s disease and dementia: a community-based cohort study. Lancet Neurol 2018; 17(5): 434–444.

57. Longford NT. Classification in two-stage screening. Stat Med 2015; 34(25): 3281–3297.

58. Sleegers K, Bettens K, De Roeck A, Van Cauwenberghe C, Cuyvers E, Verheijen J et al. A 22-single nucleotide polymorphism Alzheimer’s disease risk score correlates with family history, onset age, and cerebrospinal fluid Abeta42. Alzheimers Dement 2015; 11(12): 1452–1460.

59. Marioni RE, Campbell A, Hagenaars SP, Nagy R, Amador C, Hayward C et al. Genetic stratification to identify risk groups for Alzheimer’s disease. J Alzheimers Dis 2017; 57(1): 275–283.

60. Bellou E, Baker E, Leonenko G, Bracher-Smith M, Daunt P, Menzies G et al. Age-dependent effect of APOE and polygenic component on Alzheimer’s disease. Neurobiol Aging 2020; 93: 69–77.

61. Stocker H, Perna L, Weigl K, Mollers T, Schottker B, Thomsen H et al. Prediction of clinical diagnosis of Alzheimer’s disease, vascular, mixed, and all-cause dementia by a polygenic risk score and APOE status in a community-based cohort prospectively followed over 17 years. Mol Psychiatry 2020.

62. Fulton-Howard B, Goate AM, Adelson RP, Koppel J, Gordon ML, Alzheimer’s Disease Genetics C et al. Greater effect of polygenic risk score for Alzheimer’s disease among younger cases who are apolipoprotein E-epsilon4 carriers. Neurobiol Aging 2021; 99: 101 e101–101 e109.

63. Arboleda-Velasquez JF, Lopera F, O’Hare M, Delgado-Tirado S, Marino C, Chmielewska N et al. Resistance to autosomal dominant Alzheimer’s disease in an APOE3 Christchurch homozygote: a case report. Nat Med 2019; 25(11): 1680–1683.

64. Leonenko G, Baker E, Stevenson-Hoare J, Sierksma A, Fiers M, Williams J et al. Identifying individuals with high risk of Alzheimer’s disease using polygenic risk scores. Nat Commun 2021; 12(1): 4506.

65. Ware EB, Faul JD, Mitchell CM, Bakulski KM. Considering the APOE locus in Alzheimer’s disease polygenic scores in the Health and Retirement Study: a longitudinal panel study. BMC Med Genomics 2020; 13(1): 164.

66. Mukherjee S, Kim S, Gibbons LE, Nho K, Risacher SL, Glymour MM et al. Genetic architecture of resilience of executive functioning. Brain Imaging Behav 2012; 6(4): 621–633.

67. Mukherjee S, Kim S, Ramanan VK, Gibbons LE, Nho K, Glymour MM et al. Gene-based GWAS and biological pathway analysis of the resilience of executive functioning. Brain Imaging Behav 2014; 8(1): 110–118.

68. Hohman TJ, Dumitrescu L, Cox NJ, Jefferson AL, Alzheimer’s Neuroimaging I. Genetic resilience to amyloid related cognitive decline. Brain Imaging Behav 2017; 11(2): 401–409.

69. Felsky D, Xu J, Chibnik LB, Schneider JA, Knight J, Kennedy JL et al. Genetic epistasis regulates amyloid deposition in resilient aging. Alzheimers Dement 2017; 13(10): 1107–1116.

70. Ridge PG, Karch CM, Hsu S, Arano I, Teerlink CC, Ebbert MTW et al. Linkage, whole genome sequence, and biological data implicate variants in RAB10 in Alzheimer’s disease resilience. Genome Med 2017; 9(1): 100.

