## Supplementary Material for "Polygenic resilience scores capture protective genetic effects for Alzheimer’s disease"

### 1. **Supplementary methods**

#### **1.1 Study description**

All 26 studies in the discovery stage came from the Alzheimer’s disease (AD) genome-wide association study (GWAS) meta-analysis stage 1 published by the International Genomics of Alzheimer’s Project (IGAP)^1^, which included samples from four consortia: the Alzheimer's Disease Genetics Consortium (ADGC)^2^, the Cohorts for Heart and Aging Research in Genomic Epidemiology (CHARGE)^3^, the European Alzheimer’s disease Initiative (EADI)^4^, and the Genetic and Environmental Risk in Alzheimer’s Disease (GERAD)^5^. We acquired 21 studies from ADGC in our analyses. Two studies from CHARGE were included in our work, namely the Cardiovascular Health Study (CHS) and the Framingham Heart Study (FHS). Studies in EADI or GEARD were combined and analyzed as a single study.

Eight studies were collected to replicate polygenic resilience scores, which comprise participants from the AddNeuroMed consortium, the Alzheimer’s Disease Center wave 7 (ADC7) study of the Alzheimer’s Disease Genetics Consortium (ADGC), the Alzheimer's Disease Neuroimaging Initiative (ADNI), the Alzheimer workgroup initiative of the Psychiatric Genomic Consortium (PGC-ALZ), the Australian Imaging, Biomarker & Lifestyle Study (AIBL), and the Sydney Memory and Ageing Study (Sydney MAS). Five studies from PGC-ALZ provided data for this analysis, including the Norwegian DemGene network, the TwinGene, the Aging in Women and Men (GENDER), the Swedish Adoption/Twin Study of Aging (SATSA), and the Study of Dementia in Swedish Twins (HARMONY). Participants from GENDER, SATSA, and HARMONY studies were merged and analyzed as a single study. The eight replication studies are fully independent of the 26 studies in the discovery stage. Detailed descriptions for AddNeuroMed, ADC7, PGC-ALZ, AIBL, and Sydney MAS were published previously^1, 6-9^.

ADNI data used in the preparation of this article were obtained from the Alzheimer’s Disease Neuroimaging Initiative database (adni.loni.usc.edu). The ADNI was launched in 2003 as a public-private partnership, led by Principal Investigator Michael W. Weiner, MD. The primary goal of ADNI has been to test whether serial magnetic resonance imaging (MRI), positron emission tomography (PET), other biological markers, and clinical and neuropsychological assessment can be combined to measure the progression of mild cognitive impairment (MCI) and early Alzheimer’s disease (AD).The combined data consisting of three ADNI phases (ADNI-GO/2/3) were used as a single replication study, which has non-overlapping samples with ADNI phase 1 study (ADNI-1) in the discovery stage. Further details of ADNI study designs can be found on <http://adni.loni.usc.edu>.

Phenotypes and pre-imputation genotypes of ADGC, ADNI, and AddNeuroMed studies were downloaded for analyses in local computer. Resilience GWAS summary statistics generated on CHARGE, EADI, and GERAD samples were shared with us. Summary statistics of polygenic resilience scores in replicating the association with resilience were obtained from PGC-ALZ, AIBL, and Sydney MAS.

Notably, we used 60 years old rather than well recognized 65 years old as the age cutoff for late-onset Alzheimer’s disease (LOAD), considering the most common criterion for inclusion in the locally available LOAD GWAS studies is 60 (18 out of the 24 studies). In the current analysis, all AD cases and normal controls that had age-at-onset (AAO)/age-at-last-examination (AAE) less than 60 were excluded. Participants included in the current work were of European ancestry. Cases had clinical or autopsy-confirmed Alzheimer’s-type dementia according to NINCDS-ADRDA (National Institute of Neurological and Communicative Diseases and Stroke/Alzheimer's Disease and Related Disorders Association) or the DSM (Diagnostic and Statistical Manual of Mental Disorders, version III-V) criteria. Controls did not meet NINCDS-ADRDA/DSM III-V criteria for dementia as of the most recent follow-up.

#### **1.2 Samples and genotypes quality control**

##### **ADGC, ADNI, AddNeuroMed**

In locally downloaded studies, controls with a diagnosis of mild cognitive impairment were excluded from our analyses. Genotypes were imputed on the Michigan Imputation Server (<https://imputationserver.sph.umich.edu>) using the Haplotype Reference Consortium (HRC) r1.1 2016 reference panel^10^. Standard quality control (QC) procedures were performed on variants and samples before and after imputation using *Plink* (version 1.9: <https://www.cog-genomics.org/plink/1.9>)^11^. Call rates less than 98% were applied for both variants and samples. Common variants with minor allele frequency (MAF) ≥1% that had an imputation quality score < 0.8 were excluded from genome-wide association analysis. Furthermore, variants with Hardy–Weinberg equilibrium (HWE) *p*<1e-06 were excluded. Biallelic variants were considered for further analysis. The recorded sex of samples was compared with imputed sex, in which samples with X chromosome inbreeding coefficients <0.2 were called as female and >0.8 were called as male. Samples with conflicted sex assignments were excluded. In addition, samples with a higher degree of relatedness (PI_HAT≥0.2) were removed. Samples with any of the top 10 principal components (PCs) being more than six standard deviations away from the mean were identified as outliers and were excluded from further analysis. Variants rsIDs were obtained from dbSNP (<https://www.ncbi.nlm.nih.gov/snp/>, build 151 of genome reference GRCh37/hg19).

The post-imputation QC procedures applied in locally analyzed studies (regarding call rates, MAF, HWE, sex concordance, and sample outliers) were shared with studies that were analyzed remotely. QC procedures were substantively comparable across all studies. Additional study-specific QC steps in remotely accessible data are described below.

##### **CHARGE-FHS**

Genotype dosage data were imputed on HRC reference and were converted to hard-call genotypes using *Plink* (version 2.0). Then variants with an imputation quality score <0.8 were excluded. Family data were processed to remove samples with higher degree of relatedness (PI_HAT≥0.2).

##### **CHARGE-CHS**

Analyses were restricted to participants of European ancestry. Participants were excluded from the GWAS if they had coronary heart disease, congestive heart failure, peripheral vascular disease, valvular heart disease, stroke or transient ischemic attack or lack of available DNA at study baseline. Beyond laboratory genotyping failures, participants were excluded if they had a call rate ≤95% or if their genotype was discordant with known sex prior genotyping (to identify possible sample swaps). The following exclusions were applied to identify a final set of 306,655 autosomal single nucleotide polymorphisms (SNPs): call rate <97%, HWE *p*<1e-05, more than two duplicate errors or Mendelian inconsistencies based on Centre d'Etude du Polymorphisme Humain (CEPH) trios reference, heterozygote frequency =0, and SNPs not found in HapMap. Imputation to the HRC r1.1 2016 panel was performed on the Michigan imputation server. SNPs with the allele dosage ≤0.01 were excluded.

##### **EADI**

EADI is composed of several case-control studies and one population-based cohort, the 3C study^12, 13^. Case-control studies are comprised of AD cases and cognitively normal controls across France. The population-based cohort, the 3C study, is a prospective study of the relationship between vascular factors and dementia carried out in the three French cities Bordeaux, Montpellier and Dijon. The AD status was defined based on 12 years follow-up for Dijon participants, 14-15 years follow-up for Montpellier participants and 17-18 years follow-up for Bordeaux participants. All other non-demented participants of 3C were included as controls.

Quality control before imputation: standard quality control procedures were performed on both samples and variants prior to imputation. Samples with outlying heterozygosity, missingness >5%, or a discordant clinical and genetic gender as estimated by *Plink* (version 1.9) were excluded. Sample outliers identified by PC computed on the 1000 Genomes Phase3 reference^14^ were removed. Relatedness between samples was estimated using the *GENESIS* Package^15^ and all pairs with a kinship >0.09375 (representing the mean between the 2^nd^ and 3^rd^ degrees) were processed to remove related samples. Variants with a MAF<0.01 or with a HWE *p*<1e-6 in controls were removed. Variants with a MAF≥0.05 showing a missingness >0.05 and variants with a MAF<0.05 showing a missingness >0.02 were excluded. Finally, ambiguous variants (i.e., A/T and C/G variants) with a MAF>0.4 were also excluded.

Imputation: samples and variants passing the quality-control were used to perform the imputation with the HRC panel (Version r1.1 2016) in the Michigan Imputation Server using SHAPEIT^16^ and Minimac3^10^.

Quality control after imputation: imputed variants were converted into hard-call genotypes excluding all genotype calls with a genotype probability value below 0.8. Multi-allelic variants or variants showing an imputation quality *R^2^*<0.8 or a missingness >0.02 or a HWE *p*-value in controls <1e-6 were excluded from the analyses. The polygenic risk scores (PRSs) were computed per individual using a final set of 68,240 variants passing all the quality-control steps. The lower number of included variants in the PRS for EADI can be explained by the variants missingness resulted from the conversion of genotype dosages to hard-call genotypes and the stringent variant quality-control.

##### **GERAD**

Young population controls (<60 years old) were excluded, then sub-studies were combined and analyzed as a single study. The previous GWAS analysis of GERAD^5^ applied a covariate that combined genotyping platform and country of origin, which was not adopted in this analysis due to collinearity issues with nearly all the resilient controls came from the same country/genotyping platform. Analyses were applied to exclude SNPs showing genotype platform differences, and the PCs did not show evidence for inter-country stratification.

##### **PGC-ALZ, AIBL, Sydney MAS**

In the Norwegian DemGene network, we excluded all relatives up to 3rd degree (PI_HAT>0.125). For TwinGene study, one member of each twin pair was randomly picked for this analysis. Sydney MAS samples were genotyped using the Affymetrix Genome-wide Human SNP Array 6.0. Genotyped SNPs were excluded if: (i) the call rate was <95%, (ii) *p*-value for HWE was <10-6; (iii) minor allele frequency was <0.01 and/or (iv) if the strand was ambiguous (A/T and C/G). If first or second-degree relatives were identified, only one family member was retained for analysis. EIGENSTRAT analysis allowed for the detection and removal of any ethnic outliers. Genotypes were imputed in the Michigan imputation server using the Haplotype Reference Consortium reference panel (v3.20101123), and SNPs with high quality (imputation quality score > 0.6) were retained for analyses.

#### **1.3 HRC genome reference**

To improve the scoring performance in replication studies, we decided to choose an HRC imputed replication study instead of a discovery study as the HRC genome reference. During the stage of deriving risk score weights and preparing for resilience scripts for remotely available studies, we had access to three locally available replication datasets: ADC7, ADNI_GO2, and ADNI_GO_2. Among them, the HRC-imputed ADNI_GO2 study (original genotype file name: ADNI_GO2_GWAS_2nd_orig_BIN_I, ADNI_GO2 for short) was chosen as more common variants were preserved in this dataset by examining post-imputation QC files. In future analysis, other studies with bigger sample sizes can be used as the genome reference.

#### **1.4 Weights for polygenic risk scoring**

We obtained AD GWAS meta-analysis stage-1 summary statistics from NIAGADS (<https://www.niagads.org/home>, accession number: NG00075)^1^. We retained biallelic SNPs with MAF≥5% for risk scoring. The GWAS summary statistics were clumped (*Plink* command: --clump-kb 500 --clump-r2 0.1 --clump-p1 1.0 --clump-p2 1.0) for generating linkage-disequilibrium (LD)-independent SNPs. The effect-size weights of the risk alleles in these relatively independent SNPs were used for computing polygenic risk scores per individual. There were 160,230 independent SNPs for polygenic risk scoring in design 1. After removing *APOE* and its flanking region (chr19: 44,400kb–46,500kb), 160,082 independent SNPs remained for polygenic risk scoring in design 2.

#### **1.5 Derivation of polygenic resilience scores**

GWASs of resilience were performed using logistic regression using *Plink* (version 1.9), where the high-risk “resilient” normal controls were coded as 1. For each study, the first four PCs of ancestry were retained, along with any other of the top 20 PCs (top 10 for CHARGE-CHS study) that were significantly different between high-risk “resilient” normal controls and risk-matched late-onset Alzheimer’s disease (LOAD) cases (*p*<0.05). These PCs were used in the logistic regression model to adjust for population stratification. Additionally, AAO/AAE, and sex were used as covariates. The study site was used as a covariate for CHARGE-CHS. SNP effects on resilience across discovery-stage samples were pooled at the study-level. For both designs, we had direct access to individual participant-level data on all ADGC and ADNI samples, but only summary-level resilience GWAS results from CHARGE, EADI, and GERAD. A GWAS meta-analysis was conducted in *METAL*^17^ software using an inverse-variance random-effect model with genomic control.

In accord with the original polygenic resilience scoring pipeline described by Hess *et al*.^18^, SNPs known to be associated with LOAD risk were excluded from the resilience-scoring algorithm. Specifically, variants that showed an association with LOAD risk from the GWAS meta-analysis summary statistics^1^ (*p*<0.5), and variants that were in LD (*r^2^*≥0.2 in a 1Mb window) with those risk variants, were excluded from consideration as resilience loci. This pruning step serves as a conservative measure to avoid re-discovering risk variants for resilience scoring. Thus, the derived polygenic resilience scores were relatively independent of the polygenic risk scores that were used to stratify the high-risk samples.

For both resilience designs, the polygenic resilience score weights were generated following the same series of steps. Risk-residual SNPs from the resilience GWAS meta-analysis summary statistics were examined according to the following exclusion criteria recommended by the schizophrenia working group of the Psychiatric Genomics Consortium^19^ to retain informative SNPs for scoring: 1) variants with study-wise MAF≥5% were considered, 2) variants in the major histocompatibility complex (MHC) region (chr6:25Mb-34Mb) and chromosome 8 inversion region (chr8:7Mb-14Mb) were removed, 3) SNPs with strand-ambiguous major and minor alleles (A/T or G/C) were excluded, 4) insertion/deletion variants and SNPs sharing the same genomic position were filtered out. Additionally, we removed variants present in fewer than five studies (i.e., in <20% of the discovery studies). Relatively independent SNPs were selected using LD-clumping (*Plink* command: --clump-kb 250 --clump-r2 0.2 --clump-p1 1.0 --clump-p2 1.0).

Polygenic resilience scores were derived in a manner similar to the polygenic risk-scoring algorithm^19^. First, we counted the number of protective alleles of each SNP for each resilient sample (i.e., 0, 1, 2). Next, we derived the weighted allele counts by multiplying the allele counts with the weights for each SNP, where the weights were the natural logarithm of odds ratio (OR) of SNPs associated with resilience obtained from the resilience GWAS. The polygenic resilience score for a resilient sample was quantified by summing up the weighted allele counts throughout the genome. Multiple polygenic resilience scores can be calculated within sets of the independent marginal SNPs defined by association *p*-value cutoffs. In our analysis, we computed polygenic resilience scores within 10 *p*-value thresholds (i.e., 0.0005, 0.001, 0.01, 0.05, 0.1, 0.2, 0.3, 0.4, 0.5, 1) using *Plink* (version 1.9).

Due to the study-specific differences in genotype quality, a range of SNPs were mapped between post-imputation-QC genotypes and the list of SNPs for calculating polygenic risk scores and polygenic resilience scores. See **Supplementary Table 4** for the number of SNPs counted in risk scoring and resilience scoring in each study.

#### **1.6 Statistical analysis of resilience scores**

We estimated the variance in resilience that can be accounted for by polygenic resilience scores using two logistic regression models^19^ in independent replication studies. A full model was built (resilience status ~ polygenic resilience score + covariates) for each dataset, where the polygenic resilience score was one of the predictors for resilience status, in addition to covariates (AAO/AAE, sex, and PCs as described above). The odds of being a high-risk “resilient” normal control versus a risk-matched LOAD case per standard unit increase of polygenic resilience score were estimated. A second reduced logistic regression model (resilience status ~ covariates) was performed to estimate the amount of additive variance of resilience status contributed by the covariates as specified in the full regression model. The variance in resilience in each model was converted to Nagelkerke’s pseudo-*R^2^* using the *R* package *fmsb*. The difference in *R^2^* between the full and reduced models was computed to measure the variance of resilience status that was explained by polygenic resilience scores. The proportion of variance explained in the liability scale was derived from Nagelkerke’s pseudo-*R^2^*.^20^ For design 1, the baseline populational resilience prevalence of 10% was assumed based on the 90% cutoff used to define high-risk “resilient” normal controls. For design 2, the baseline population resilience prevalence of 2.8% was estimated jointly based on the *APOE*-ε4 prevalence in the European ancestry population (14%)^21, 22^ and the high-PRS cutoff (80%). A meta-analysis of the natural logarithm of OR and standard error of polygenic resilience scores was conducted using the *R* package *metafor* with an inverse-variance random-effects model. The variances explained by polygenic resilience scores were pooled across replication studies using a weighted mean, where the weight for each study was from the meta-analysis model.

Three of the eight replication studies (ADC7, AddNeuroMed, and ADNI-GO/2/3; see **Table 1** and **Supplementary Table 1**) were directly accessible by downloading onto local computers and were used for performing a series of correlation analyses for polygenic resilience scores. Firstly, the Pearson’s correlation coefficients were calculated for examining the correlation of polygenic risk and resilience scores in separate groups: (1) all LOAD cases; (2) all normal controls. The correlation analysis was repeated after retaining high-risk sample groups defined by each design. In this analysis, we excluded ultra-high-risk LOAD cases whose PRSs are higher than the maximum of all normal controls, and ultra-low-risk normal controls whose PRSs are lower than the minimum of all LOAD cases. Secondly, using AAO/AAE, sex, and four PCs as covariates, logistic regression models were employed to evaluate the OR of being in a group versus the other per standard unit increase of polygenic resilience scores: (1) *APOE*-ε4 carriers in high-risk “resilient” normal controls vs. *APOE*-ε4 carriers in risk-matched LOAD cases; and (2) *APOE*-ε4 carriers in high-risk “resilient” normal controls vs. *APOE*-ε4 non-carriers in high-risk “resilient” normal controls.

### 2. **Supplementary results**

#### **2.1 Resilient sample distribution in the discovery and replication**

In the discovery stage, the ratio of high-risk “resilient” normal controls to risk-matched LOAD cases was ~1:5 (2263:11309) in design 1, and ~1:7 (988:6541) in design 2. In the replication stage, the ratio was ~3:1 (1056:381) in design 1, and ~2:1 (583:331) in design 2. The different high-risk control/case ratio in replication stage from the discovery stage can be due to two aspects. On one hand, as the studies included in the discovery stage were used to derive PRS weights, the performance of PRS in distinguishing AD cases and controls in the discovery stage reflected overfitting. Thus, we also expected that the distributions of PRS between controls and cases would be less separated in replication studies, which were fully independent of discovery studies. This phenomenon resulted in a lower percentage of the cases being retained in the resilience analysis at the replication stage. On the other hand, we had four larger studies (GENDER/SATSA/HARMONY, TwinGene, AIBL, and Sydney MAS) that sampled many more controls than cases. TwinGene accounted for ~60% of high-risk controls but produced only ~30 risk-matched cases. Thus, the difference in sample ratio was also a byproduct of the pipeline being applied to already imbalanced data. However, a disparity in high-risk control: case ratio would not necessarily be detrimental to a polygenic scoring analysis if 1) sample size was sufficiently large in the replication sample, and 2) the initial training sample and model building were optimal to yield a generalizable resilience score. Ideally, increasing the ratio of risk-matched cases, which we used as comparisons to high-risk “resilient” controls, would increase the power of replicating the association with resilience. Nonetheless, our analysis showed we had enough power to successfully replicate polygenic resilience scores under design 1.

#### **2.2 Resilience GWAS**

After pruning risk-associated SNPs, SNPs in LD with risk-associated SNPs, and QC-excluded SNPs, far fewer SNPs remained for association testing than the number typically seen for a risk GWAS. In fact, we ended up with less than 100,000 SNPs to test, whereas a typical GWAS has around 5~6 million SNPs. Across the 94,655 common variants (MAF≥5%) examined in design 1, one SNP showed association with resilience to LOAD at a suggestive level of significance (*p*<1e-05) (**Supplementary Figure 1A**), specifically, rs2194962 (*p*=6.68e-07). In GRCh37 reference genome with 0kb gene window, rs2194962 (chr11:60065105) was annotated as an intron variant, nonsense-mediated decay variant, and regulatory region variant on *MS4A4A* (Membrane-Spanning 4-Domains, Subfamily A, Member 4). Note that rs2194962 was present in only three out of 26 studies in the meta-analysis; further investigation of its role in resilience was warranted. In contrast, as anticipated, none of the 86,752 common SNPs displayed genome-wide significant association (*p*<5e-08), and none surpassed even the suggestive significance threshold (*p*<1e-05) in design 2. For both designs, we examined population stratification effects by comparing the resilience association *p*-values of the marginal SNPs against the expected uniform distribution of *p*-values. The quantile-quantile (QQ) plots revealed that the observed association *p*-values fit closely with the expected p-value distribution (**Supplementary Figure 2**). The genome inflation factor (λgc) was 0.94 and 0.97 for design 1 and design 2, respectively.

#### **2.3 Interaction of risk and resilience effects**

In locally available replication studies (ADC7, AddNeuroMed, and ADNI-GO/2/3), resilience scores from *p*<0.1 threshold in design 1 showed a nonsignificant positive correlation with risk scores among high-risk “resilient” normal controls (Pearson’s *r*=0.046, 95% confidence interval [CI]=-0.130-0.219, degree of freedom [*df*]=124, *p*=0.61), and a nonsignificant negative correlation among risk-matched LOAD cases (Pearson’s *r*=-0.032, 95% CI=-0.210-0.148, *df*=118, *p*=0.73). In high-risk “resilient” normal controls, power analysis demonstrated we didn’t have sufficient statistical power (1- β=0.2) to detect a significant correlation as strong as the correlation in all normal controls (Pearson’s *r*=0.102). In addition, we had very low power (1- β=0.09) in identifying significant differences between correlations in high-risk “resilient” normal controls and risk-matched LOAD cases. The lack of power indicates investigating the correlations between risk and resilience scores using larger high-risk samples is warranted.

In *APOE*-ε4 carriers of locally available replication studies, we found increasing polygenic resilience scores were significantly associated with higher odds of being a high-risk “resilient” normal control than a risk-matched LOAD (*p*<0.1 threshold of design 1: OR=1.64, 95% CI=1.08-2.50, *p*=0.021; *p*<0.2 threshold of design 1: OR=1.98, 95% CI=1.24-3.15, *p*=3.9e-03). Among high-risk “resilient” normal controls, we observed a significant positive association of resilience scores (*p*<0.2 threshold of design 1) with the odds of carrying *APOE*-ε4 allele(s) relative to those not carrying *APOE*-ε4 allele(s) (OR=1.57, 95% CI=1.07-2.29, *p*=0.021).

### 3. **Supplementary Figures**

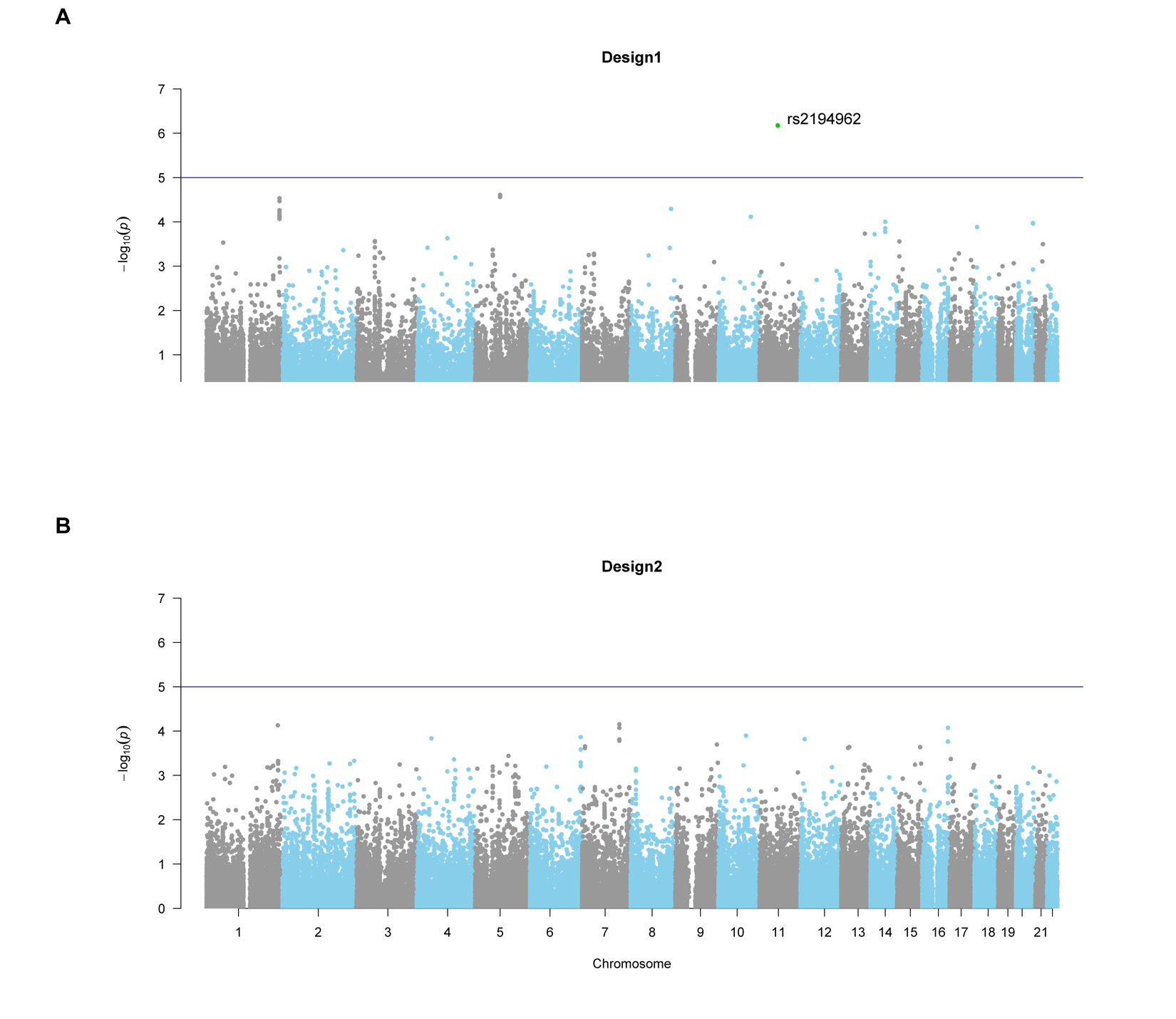

**Supplementary Figure 1**. **Manhattan plots showing genome-wide association study (GWAS) meta-analysis results for resilience to late-onset Alzheimer’s disease (A, design 1; B, design 2).** The resilience-association *p*-values (y-axis) for risk-independent common single nucleotide polymorphisms (SNPs, minor allele frequency ≥5%) in all 22 autosomes (x-axis) are represented as dots. Panel A shows 94,655 SNPs for design 1 and panel B shows 86,752 SNPs for design 2. The horizontal blue line denotes a “suggestive” level of association for GWAS (i.e., *p*≤1.0e-05), and green dot labelled with rsID is SNP above this suggestive significance threshold. As expected, no SNPs in either design attained genome-wide significant association due to attenuated sample size.

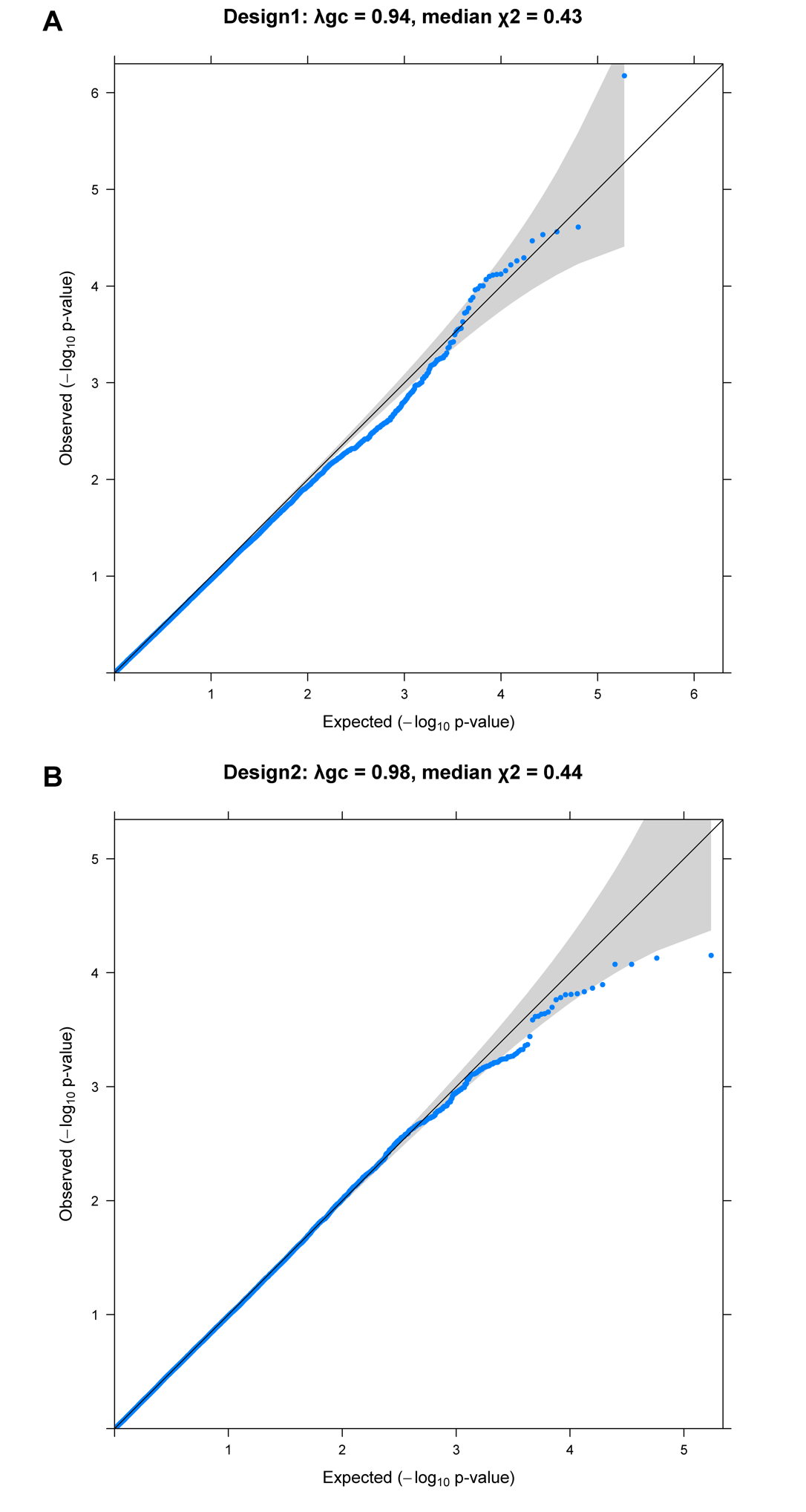

**Supplementary Figure 2. Quantile-quantile (QQ) plots (A, design 1; B, design 2).** The quantiles of observed *p*-values from the genome-wide association study of resilience to late-onset Alzheimer’s disease (y-axis) are plotted against the quantiles of expected *p*-values from a uniform distribution (x-axis). The 95% confidence intervals of the expected distributions of *p*-values are shown by grey shading.

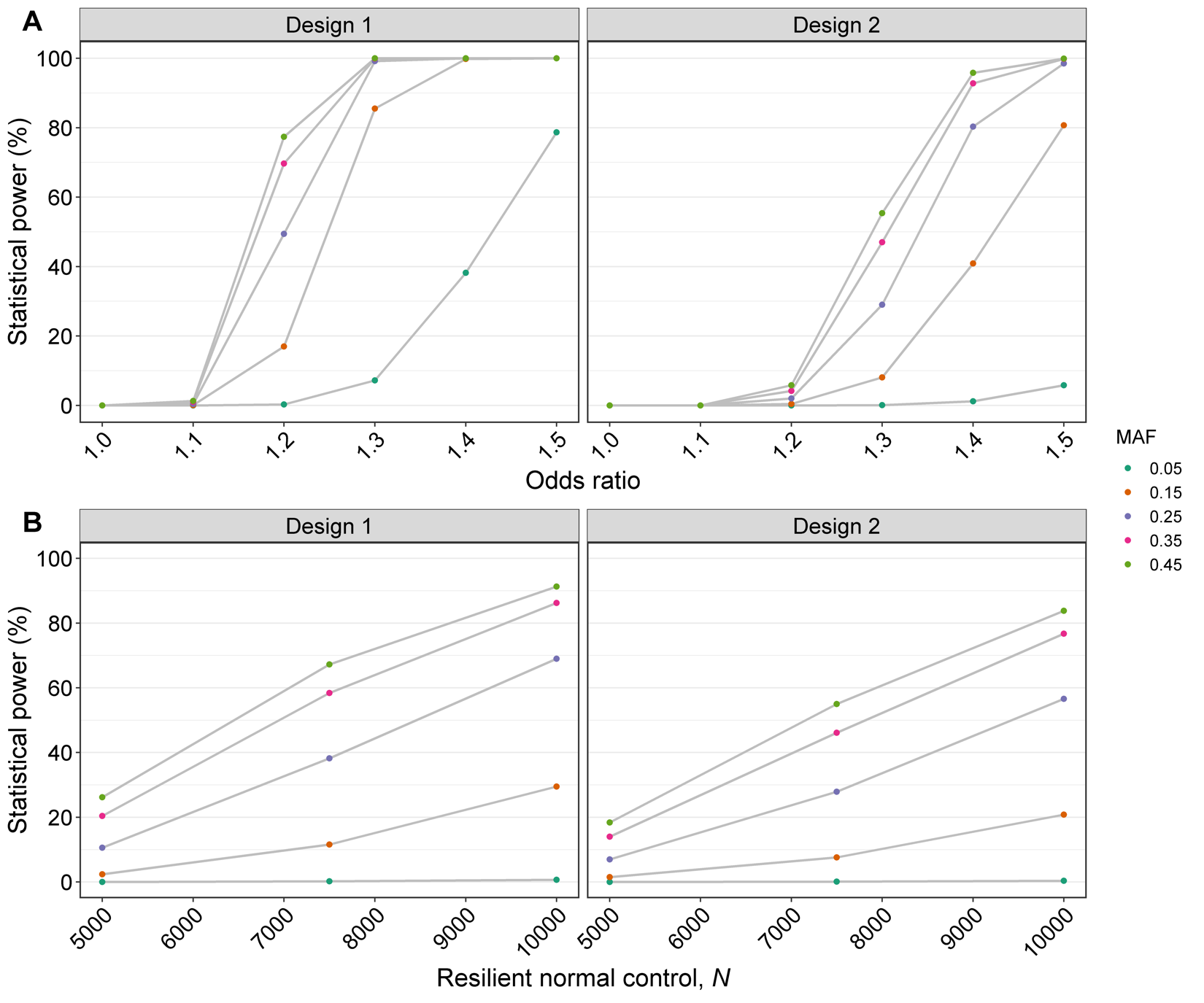

**Supplementary Figure 3.** **Estimated** **statistical power of resilience genome-wide association study (GWAS)** **(left panels: design 1; right panels: design 2).** The statistical power was estimated using Genetic Association Study (GAS) Power Calculator (https://csg.sph.umich.edu/abecasis/gas_power_calculator/). The populational prevalence of resilience was assumed to be 10% in design 1, and 2.8% in design 2. Panel A demonstrates the power of identifying genome-wide significant single nucleotide polymorphisms (SNPs) (*p*<5e-08) under various scenarios of effect-size and minor allele frequency, using the sample size of “resilient” high-risk controls and risk-matched late-onset Alzheimer’s disease (LOAD) cases in our discovery studies. In panel B, considering the possibility of the winner’s curse and an over-estimation of effect sizes for resilience, we used a more reasonable effect size (odds ratio=1.1, similar to that for common variants most strongly associated with other complex disorders by GWAS) to estimate the power of identifying genome-wide significant SNPs when more samples are added. The ratio of “resilient” high-risk normal controls to risk-matched LOAD cases was assumed to be 0.2 (as seen in the current study) in design 1, and a ratio of 0.15 was applied in design 2.

**Supplementary Table 1. The number and the age-at-onset/age-at-last-examination of LOAD cases and normal controls, high-risk normal controls ("resilient" individuals) and risk-matched LOAD cases identified in each of the discovery and replication studies.**

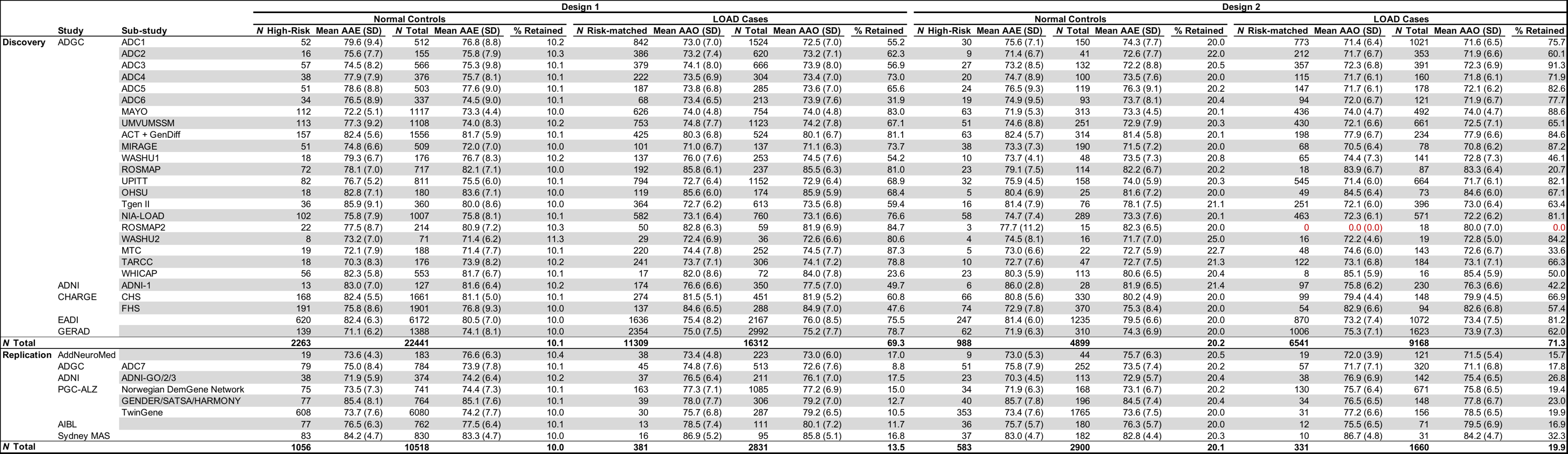

Abbreviations: LOAD, late-onset Alzheimer's disease; AAE, age-at-last-examination; AAO, age-at-onset; SD, standard deviation. A list of study full names is in Supplementary Table 2.

Note: "Retained" column indicates the percentage of high-risk normal controls of all normal controls retained for resilience genome-wide association analysis per study, or the percentage of risk-matched LOAD cases of all LOAD cases retained in analysis per study. ROSMAP2 study had no LOAD cases (highlighted in red) whose risk matched with high-risk normal controls in design 2, and was not included in analysis for design 2.

**Supplementary Table 2. The acronym, full name, and data availability of the discovery and replication studies.**

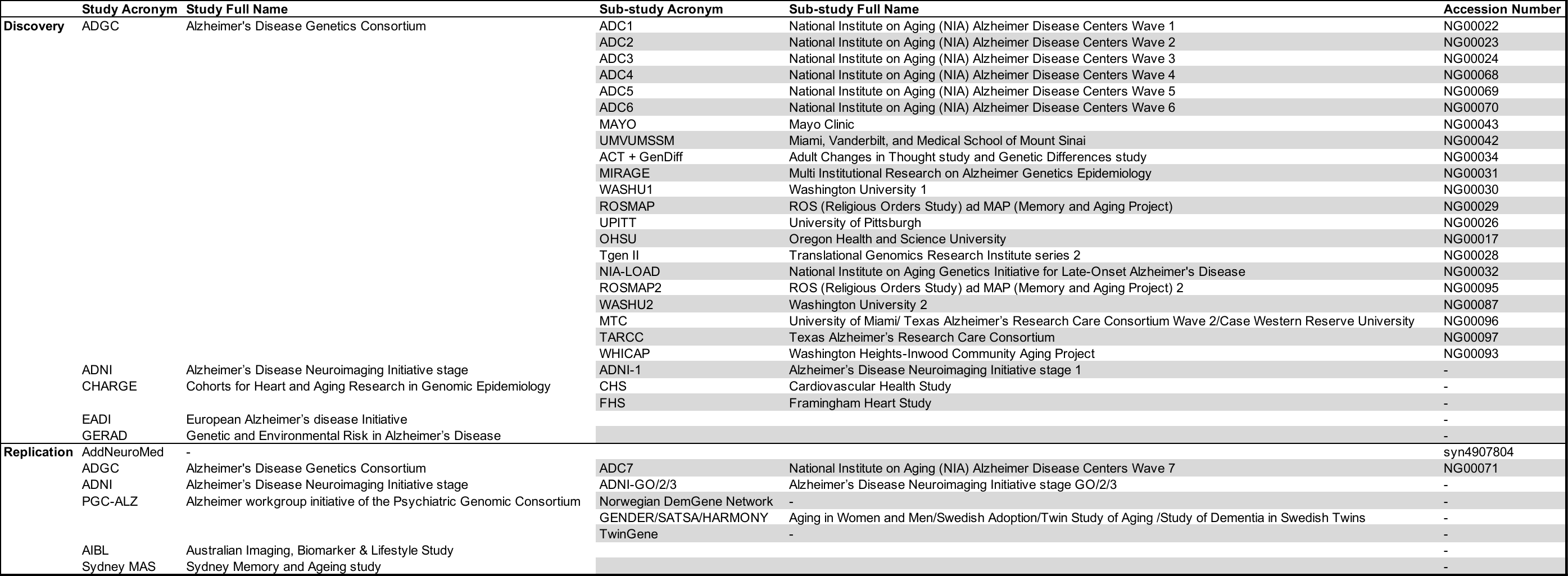

Note: The ADGC, ADNI, and AddNeuroMed data used in this study were provided under restricted access by NIAGADS (https://www.niagads.org), ADNI (http://adni.loni.usc.edu), and Synapse platform (https://www.synapse.org), respectively. The accession numbers of studies from NIAGADS and Synapse platform are listed. Only summary statistics were made available to us from EADI, GERAD, CHARGE, PGC, AIBL, and Sydney MAS.

**Supplementary Table 3. Parameters for individual steps of the analysis.**

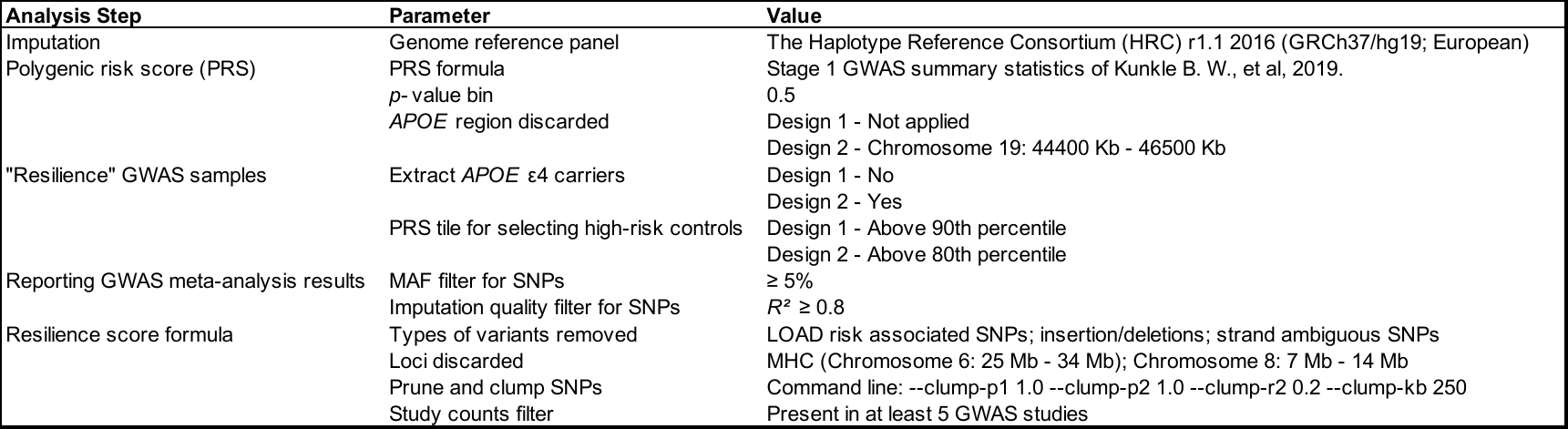

Abbreviations: LOAD, late-onset Alzheimer's disease; GWAS, genome-wide genotype data; *APOE*, apolipoprotein E; MAF, minor allele frequency; SNPs, single nucleotide polymorphisms; MHC, major histocompatibility complex.

**Supplementary Table 4. Total number of SNPs included in polygenic risk scores and polygenic resilience scores in each study.**

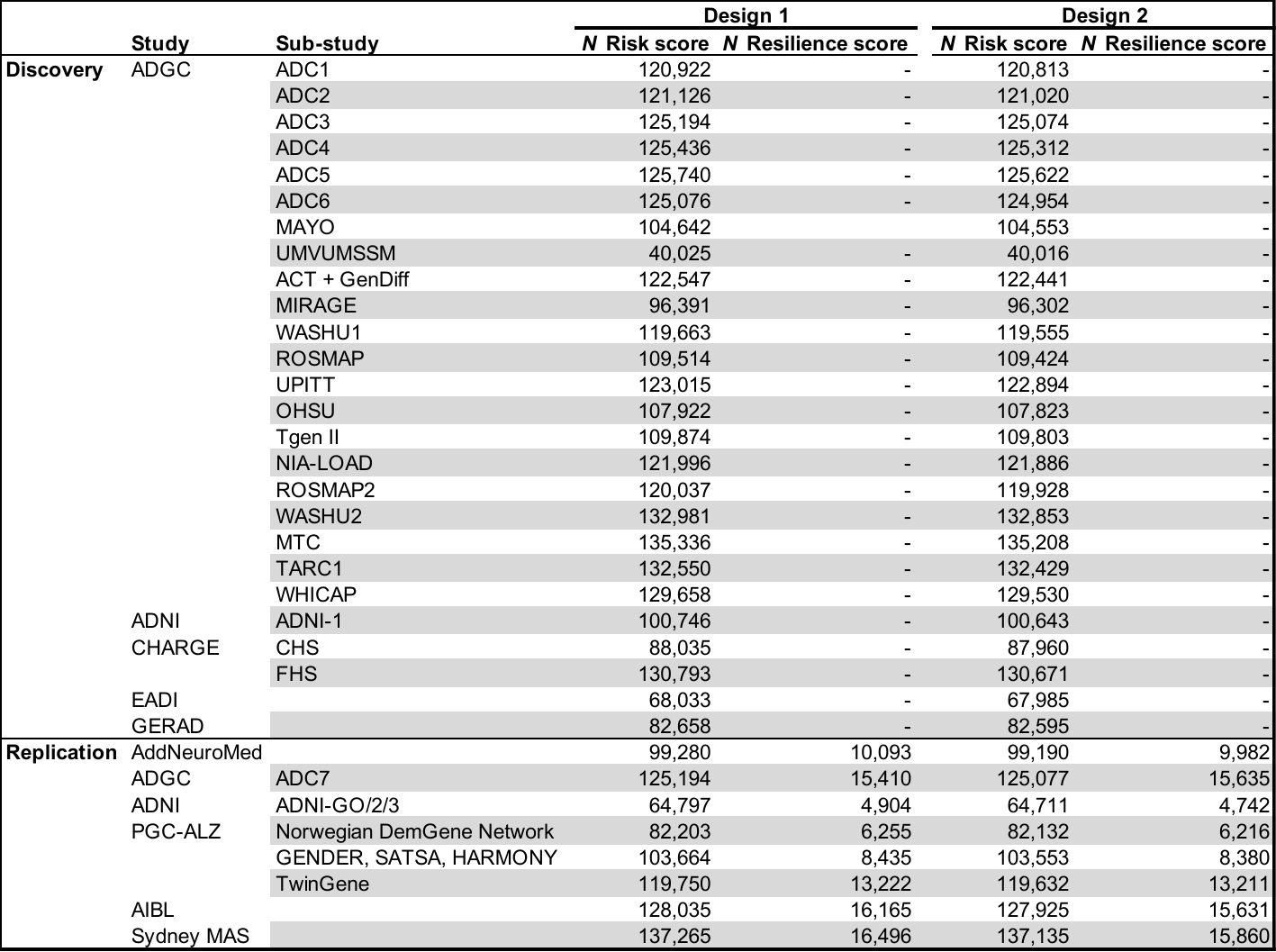

Abbreviations: SNPs = single nucleotide polymorphisms. A list of study full names is in Supplementary Table 2.

### 5. **Supplementary acknowledgements**

Stephen Glatt is supported by the U.S. National Institutes of Health (NIMH) [grant number 5R01MH101519], the U.S. National Institute on Aging (NIA) [grant numbers R01 AG064955 and 5R01AG054002], the Sidney R. Baer, Jr. Foundation, and NARSAD: The Brain & Behavior Research Foundation. William Kremen is supported by NIA [grant numbers R01 AG050595 and R01 AG022381]. Valentina Escott-Price is supported by the UK Dementia Research Institute (UKDRI, which is supported by the Medical Research Council [UKDRI-3003], Alzheimer’s Research UK, and Alzheimer’s Society), Joint Programming for Neurodegeneration [MRC: MR/T04604X/1], and Dementia Platforms UK [MRC: MR/L023784/2]. Christine Fennema-Notestine is supported by awards from the National Institutes of Health/National Institute on Aging [grant numbers R01s AG064955, AG022381, AG062483, and P01 AG055367]. Stephen Faraone is supported by the K.G. Jebsen Centre for Research on Neuropsychiatric Disorders, University of Bergen, Bergen, Norway, the European Union’s Seventh Framework Programme for research, technological development and demonstration [grant number 602805], the European Union’s Horizon 2020 research and innovation programme [grant number 667302], and NIMH [grant numbers 5R01MH101519 and U01 MH109536-01]. Simon Laws is supported by National Health and Medical Research Council (NHMRC) [grant numbers APP1161706, APP1191535 and APP1151854]. Margaret Gatz is supported by NIH [grant number R01 AG060470].

Funding for the AIBL study was provided in part by the study partners (Commonwealth Scientific Industrial and research Organization (CSIRO), Edith Cowan University (ECU), Mental Health Research institute (MHRI), National Ageing Research Institute (NARI), Austin Health, CogState Ltd.). The AIBL study has also received support from the NHMRC and the Dementia Collaborative Research Centres program (DCRC2), as well as funding from the Science and Industry Endowment Fund (SIEF) and the Cooperative Research Centre (CRC) for Mental Health—funded through the CRC Program [grant number 20100104], an Australian Government Initiative. We thank all those who took part as a participant in the AIBL study for their commitment and dedication to helping advance research into the early detection and causation of AD.

This CHS research was supported by NHLBI contracts [HHSN268201200036C, HHSN268200800007C, HHSN268201800001C, N01HC55222, N01HC85079, N01HC85080, N01HC85081, N01HC85082, N01HC85083, N01HC85086]; and NHLBI grants [grant numbers U01HL080295, R01HL087652, R01HL105756, R01HL103612, R01HL120393, and U01HL130114], with additional contribution from the National Institute of Neurological Disorders and Stroke (NINDS). Additional support was provided through the National Institute on Aging [grant numbers R01AG023629, R01AG15928, R01AG15928, and R01AG033193]. A full list of principal CHS investigators and institutions can be found at CHS-NHLBI.org. The provision of genotyping data was supported in part by the National Center for Advancing Translational Sciences, CTSI [grant number UL1TR001881], and the National Institute of Diabetes and Digestive and Kidney Disease Diabetes Research Center (DRC) [grant number DK063491] to the Southern California Diabetes Endocrinology Research Center. The content is solely the responsibility of the authors and does not necessarily represent the official views of the National Institutes of Health.

The Sydney MAS has been funded by three National Health & Medical Research Council Program Grants [grant numbers ID350833, ID568969, APP1093083]. We thank the participants and their informants for their time and generosity in contributing to this research. We also acknowledge the Sydney MAS research team: [https://cheba.unsw.edu.au/research-projects/sydney-memory-and-ageing-study](https://urldefense.com/v3/__https:/cheba.unsw.edu.au/research-projects/sydney-memory-and-ageing-study__;!!GobTDDpD7A!bNpJrdDuWd_nXconGPCrCqDn9J-BRTTy5qwVUIWESMqF5m-svHlv4ISfj5gvFsfQ$).

Data collection and sharing for this project was funded by the Alzheimer's Disease Neuroimaging Initiative (ADNI) (National Institutes of Health [grant number U01 AG024904]) and DOD ADNI (Department of Defense award number W81XWH-12-2-0012). ADNI is funded by the National Institute on Aging, the National Institute of Biomedical Imaging and Bioengineering, and through generous contributions from the following: AbbVie, Alzheimer’s Association; Alzheimer’s Drug Discovery Foundation; Araclon Biotech; BioClinica, Inc.; Biogen; Bristol-Myers Squibb Company; CereSpir, Inc.; Cogstate; Eisai Inc.; Elan Pharmaceuticals, Inc.; Eli Lilly and Company; EuroImmun; F. Hoffmann-La Roche Ltd and its affiliated company Genentech, Inc.; Fujirebio; GE Healthcare; IXICO Ltd.; Janssen Alzheimer Immunotherapy Research & Development, LLC.; Johnson & Johnson Pharmaceutical Research & Development LLC.; Lumosity; Lundbeck; Merck & Co., Inc.; Meso Scale Diagnostics, LLC.; NeuroRx Research; Neurotrack Technologies; Novartis Pharmaceuticals Corporation; Pfizer Inc.; Piramal Imaging; Servier; Takeda Pharmaceutical Company; and Transition Therapeutics. The Canadian Institutes of Health Research is providing funds to support ADNI clinical sites in Canada. Private sector contributions are facilitated by the Foundation for the National Institutes of Health (www.fnih.org). The grantee organization is the Northern California Institute for Research and Education, and the study is coordinated by the Alzheimer’s Therapeutic Research Institute at the University of Southern California. ADNI data are disseminated by the Laboratory for Neuro Imaging at the University of Southern California.

The authors would like to acknowledge the contributions of Dr. Rahul Desikan to the development of this study. Dr. Desikan, who passed away on July 14, 2019 due to complications from ALS, was a consultant on the project.

### **References**

1.	Kunkle BW, Grenier-Boley B, Sims R, Bis JC, Damotte V, Naj AC *et al.* Genetic meta-analysis of diagnosed Alzheimer's disease identifies new risk loci and implicates Abeta, tau, immunity and lipid processing. *Nat Genet* 2019; **51**(3)**:** 414-430.

2.	Naj AC, Jun G, Beecham GW, Wang LS, Vardarajan BN, Buros J *et al.* Common variants at MS4A4/MS4A6E, CD2AP, CD33 and EPHA1 are associated with late-onset Alzheimer's disease. *Nat Genet* 2011; **43**(5)**:** 436-441.

3.	Psaty BM, O'Donnell CJ, Gudnason V, Lunetta KL, Folsom AR, Rotter JI *et al.* Cohorts for Heart and Aging Research in Genomic Epidemiology (CHARGE) Consortium: Design of prospective meta-analyses of genome-wide association studies from 5 cohorts. *Circ Cardiovasc Genet* 2009; **2**(1)**:** 73-80.

4.	Lambert JC, Ibrahim-Verbaas CA, Harold D, Naj AC, Sims R, Bellenguez C *et al.* Meta-analysis of 74,046 individuals identifies 11 new susceptibility loci for Alzheimer's disease. *Nat Genet* 2013; **45**(12)**:** 1452-1458.

5.	Harold D, Abraham R, Hollingworth P, Sims R, Gerrish A, Hamshere ML *et al.* Genome-wide association study identifies variants at CLU and PICALM associated with Alzheimer's disease. *Nat Genet* 2009; **41**(10)**:** 1088-1093.

6.	Proitsi P, Lupton MK, Velayudhan L, Newhouse S, Fogh I, Tsolaki M *et al.* Genetic predisposition to increased blood cholesterol and triglyceride lipid levels and risk of Alzheimer disease: a Mendelian randomization analysis. *PLoS Med* 2014; **11**(9)**:** e1001713.

7.	Lovestone S, Francis P, Kloszewska I, Mecocci P, Simmons A, Soininen H *et al.* AddNeuroMed--the European collaboration for the discovery of novel biomarkers for Alzheimer's disease. *Ann N Y Acad Sci* 2009; **1180:** 36-46.

8.	Jansen IE, Savage JE, Watanabe K, Bryois J, Williams DM, Steinberg S *et al.* Genome-wide meta-analysis identifies new loci and functional pathways influencing Alzheimer's disease risk. *Nat Genet* 2019; **51**(3)**:** 404-413.

9.	Zhang Q, Sidorenko J, Couvy-Duchesne B, Marioni RE, Wright MJ, Goate AM *et al.* Risk prediction of late-onset Alzheimer's disease implies an oligogenic architecture. *Nat Commun* 2020; **11**(1)**:** 4799.

10.	Das S, Forer L, Schonherr S, Sidore C, Locke AE, Kwong A *et al.* Next-generation genotype imputation service and methods. *Nat Genet* 2016; **48**(10)**:** 1284-1287.

11.	Chang CC, Chow CC, Tellier LC, Vattikuti S, Purcell SM, Lee JJ. Second-generation PLINK: rising to the challenge of larger and richer datasets. *Gigascience* 2015; **4:** 7.

12.	Group CS. Vascular factors and risk of dementia: design of the Three-City Study and baseline characteristics of the study population. *Neuroepidemiology* 2003; **22**(6)**:** 316-325.

13.	Lambert JC, Heath S, Even G, Campion D, Sleegers K, Hiltunen M *et al.* Genome-wide association study identifies variants at CLU and CR1 associated with Alzheimer's disease. *Nat Genet* 2009; **41**(10)**:** 1094-1099.

14.	Genomes Project C, Auton A, Brooks LD, Durbin RM, Garrison EP, Kang HM *et al.* A global reference for human genetic variation. *Nature* 2015; **526**(7571)**:** 68-74.

15.	Gogarten SM, Sofer T, Chen H, Yu C, Brody JA, Thornton TA *et al.* Genetic association testing using the GENESIS R/Bioconductor package. *Bioinformatics* 2019; **35**(24)**:** 5346-5348.

16.	Delaneau O, Marchini J, Zagury JF. A linear complexity phasing method for thousands of genomes. *Nat Methods* 2011; **9**(2)**:** 179-181.

17.	Willer CJ, Li Y, Abecasis GR. METAL: fast and efficient meta-analysis of genomewide association scans. *Bioinformatics* 2010; **26**(17)**:** 2190-2191.

18.	Hess JL, Tylee DS, Mattheisen M, Schizophrenia Working Group of the Psychiatric Genomics C, Lundbeck Foundation Initiative for Integrative Psychiatric R, Borglum AD *et al.* A polygenic resilience score moderates the genetic risk for schizophrenia. *Mol Psychiatry* 2021; **26**(3)**:** 800-815.

19.	Schizophrenia Working Group of the Psychiatric Genomics C. Biological insights from 108 schizophrenia-associated genetic loci. *Nature* 2014; **511**(7510)**:** 421-427.

20.	Lee SH, Wray NR, Goddard ME, Visscher PM. Estimating missing heritability for disease from genome-wide association studies. *Am J Hum Genet* 2011; **88**(3)**:** 294-305.

21.	Farrer LA, Cupples LA, Haines JL, Hyman B, Kukull WA, Mayeux R *et al.* Effects of age, sex, and ethnicity on the association between apolipoprotein E genotype and Alzheimer disease. A meta-analysis. APOE and Alzheimer Disease Meta Analysis Consortium. *JAMA* 1997; **278**(16)**:** 1349-1356.

22.	Bang OY, Kwak YT, Joo IS, Huh K. Important link between dementia subtype and apolipoprotein E: a meta-analysis. *Yonsei Med J* 2003; **44**(3)**:** 401-413.
